# Integrated multi-omics reveals anaplerotic insufficiency in methylmalonyl-CoA mutase deficiency

**DOI:** 10.1101/2022.01.27.22269972

**Authors:** Patrick Forny, Ximena Bonilla, David Lamparter, Wenguang Shao, Tanja Plessl, Caroline Frei, Anna Bingisser, Sandra Goetze, Audrey van Drogen, Keith Harshman, Patrick G. A. Pedrioli, Cedric Howald, Martin Poms, Florian Traversi, Sarah Cherkaoui, Raphael J. Morscher, Luke Simmons, Merima Forny, Ioannis Xenarios, Ruedi Aebersold, Nicola Zamboni, Gunnar Raetsch, Emmanouil Dermitzakis, Bernd Wollscheid, Matthias R. Baumgartner, D. Sean Froese

**Author notes:** These authors contributed equally to this study (Patrick Forny, Ximena Bonilla, David Lamparter, Wenguang Shao). These authors are listed alphabetically (Ximena Bonilla, David Lamparter, Wenguang Shao).

## Abstract

Multi-layered omics approaches can help define relationships between genetic factors, biochemical processes and phenotypes thus extending research of inherited diseases beyond identifying their monogenic cause ^1^. We implemented a multi-layered omics approach for the inherited metabolic disorder methylmalonic aciduria (MMA). We performed whole genome sequencing, transcriptomic sequencing, and mass spectrometry-based proteotyping from matched primary fibroblast samples of 230 individuals (210 affected, 20 controls) and related the molecular data to 105 phenotypic features. Integrative analysis identified a molecular diagnosis for 84% (177/210) of affected individuals, the majority (148) of whom had pathogenic variants in methylmalonyl-CoA mutase (*MMUT*). Untargeted analysis of all three omics layers revealed dysregulation of the TCA cycle and surrounding metabolic pathways, a finding that was further corroborated by multi-organ metabolomics of a hemizygous *Mmut* mouse model. Integration of phenotypic disease severity indicated downregulation of oxoglutarate dehydrogenase and upregulation of glutamate dehydrogenase, two proteins involved in glutamine anaplerosis of the TCA cycle. The relevance of disturbances in this pathway was supported by metabolomics and isotope tracing studies which showed decreased glutamine-derived anaplerosis in MMA. We further identified MMUT to physically interact with both, oxoglutarate dehydrogenase complex components and glutamate dehydrogenase providing evidence for a multi-protein metabolon that orchestrates TCA cycle anaplerosis. This study emphasizes the utility of a multi-modal omics approach to investigate metabolic diseases and highlights glutamine anaplerosis as a potential therapeutic intervention point in MMA.

**Take home message:** Combination of integrative multi-omics technologies with clinical and biochemical features leads to an increased diagnostic rate compared to genome sequencing alone and identifies anaplerotic rewiring as a targetable feature of the rare inborn error of metabolism methylmalonic aciduria.

**Graphical abstract:** 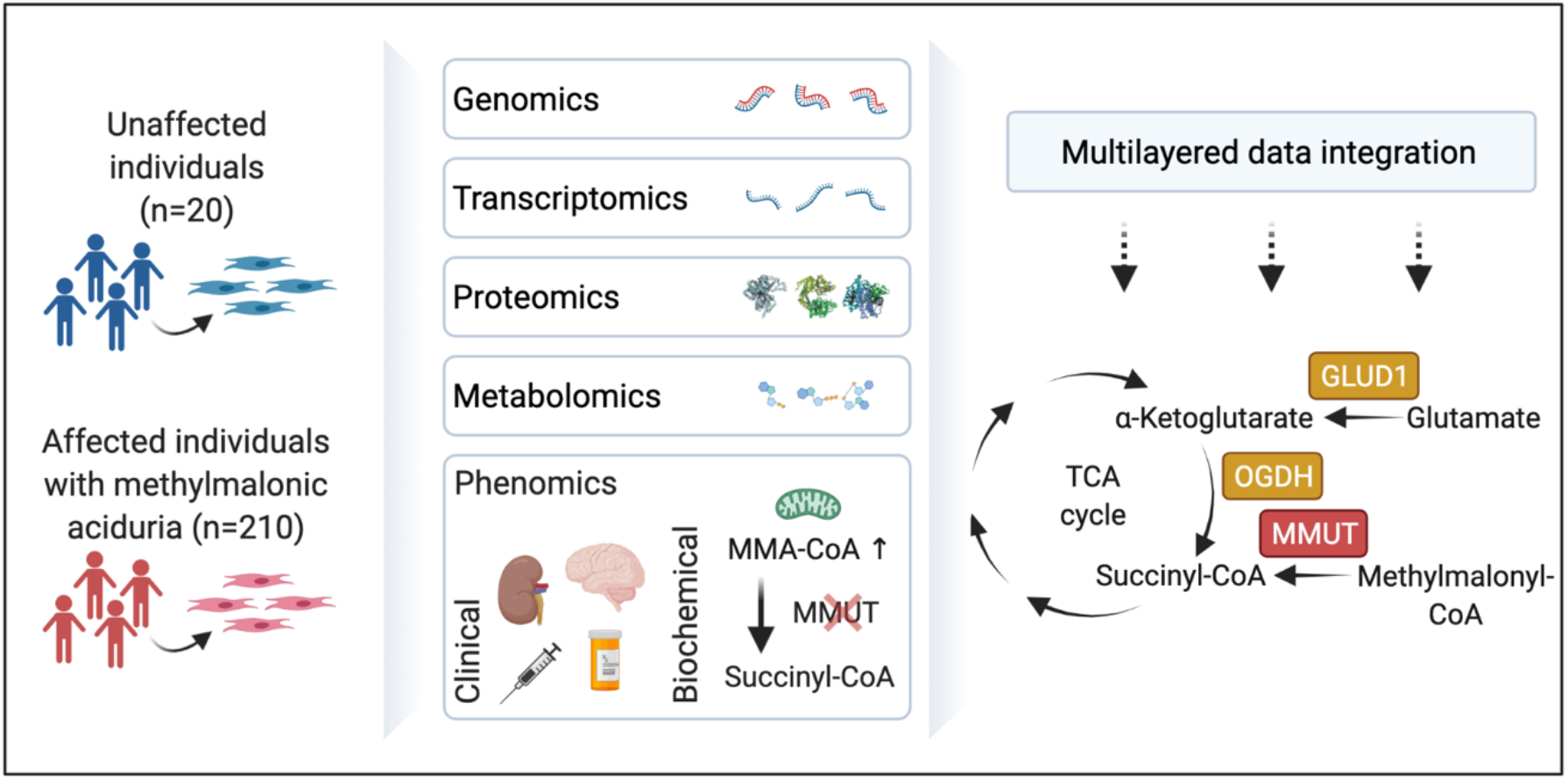

## Main

Inborn errors of metabolism (IEMs), first described by Archibald Garrod ^2^, are inherited diseases resulting in inadequate function of metabolic proteins. IEMs represent a group of nearly 1,500 diseases with a combined incidence of approximately 1:800 births. They present a clinically and genetically heterogeneous picture making them inherently difficult to diagnose ^3,4^. Beyond their diagnostic challenges, the pathomechanisms of many IEMs are not well understood, hence most IEMs lack rationalized treatment approaches ^5^.

Technological advances in genomics and mass spectrometry, leveraging datasets of whole molecule classes (omics), have recently led to a paradigm shift in their use as diagnostic tools. For example, single-layer whole genome sequencing has achieved diagnostic rates of 30-50% in rare disease cohorts ^6–8^, while dual-layer combination with RNA sequencing can improve this by 10-35% ^9–12^. In spite of these advances a significant number of patients remain undiagnosed and disease course prediction remains poor, mainly due to a lack of pathomechanistic understanding and often unclear genotype-phenotype relationships.

Multi-layered omics data have the potential to not only increase diagnosis rates of inborn errors of metabolism, but also to uncover novel mechanistic insights into disease pathophysiology ^1^, thus potentially indicating new therapeutic targets. Such a combinatorial approach is key to move beyond the traditional “one gene – one disease” view of these disorders which fails to explain phenotypic heterogeneity based on genetic variation only. However, simultaneous application of multi-omics technologies for this purpose have not been rigorously tested, and their true utility as well as bottlenecks and knowledge gaps to using them in this regard remain unknown.

Here, we have generated multi-layered omics information from individuals suspected with the prototypic IEM methylmalonic aciduria (MMA). MMA has approximately 20 potential monogenic causes most of which result in deficiency of the methylmalonyl-CoA mutase (MMUT) enzyme due to loss of function or proper cofactor (cobalamin) assembly ^13^. Even though the (dys)function of MMUT has been studied extensively ^14–17^, the main metabolic disturbances in MMA remain an open question.

By combining whole genome sequencing (WGS), whole transcriptome sequencing (RNA-seq) and proteotyping information (DIA-MS) with phenotypic features we identified disease causative and pathogenic features in a cohort of MMA affected individuals. We revealed underlying damaging variants and differentially expressed transcripts and proteins directly related to anaplerosis of the TCA cycle. Moreover, follow-up studies utilizing untargeted metabolomics and [U-^13^C]glutamine tracing revealed a depletion of TCA cycle metabolites and decreased oxidation of glutamine-derived carbons in line with the identified dysregulation of glutamate dehydrogenase and oxoglutarate dehydrogenase enzymes, which we found to physically interact with MMUT. Beyond unveiling these metabolic disturbances in MMUT deficiency, our findings enable a better biological understanding of TCA cycle anaplerosis. Furthermore, the anaplerotic TCA cycle insufficiency may be a potential intervention point in MMA to therapeutically compensate for the loss of TCA cycle intermediates.

## Results

### Monogenic disease variant detection through multi-omics

To extend the understanding of MMA from the causative genomic lesions to the affected biochemical processes, we performed high-quality WGS, RNA-seq and DIA-MS based proteotyping on fibroblasts taken from 230 individuals (210 affected by MMA, 20 unaffected), representing a mainly European cohort collected over a 25-year period (**Fig. 1a** and **Extended data Fig. 1**). Classical MMA is caused by defective processing of methylmalonyl-CoA in the catabolism of propionate, resulting from deficiency of the enzyme methylmalonyl-CoA mutase (MMUT) or proteins related to production of its cofactor adenosylcobalamin (e.g., MMAA, MMAB) or substrate (**Fig. 1a**). Biochemical assay of propionate incorporation (PI) ^18^, and MMUT enzyme activity ^14^ strongly correlated across all samples (rho=0.73, p<0.0001) (**Extended data Fig. 2a**). Fibroblasts from 150 individuals with MMA had reduced MMUT activity (MMUT-deficient), including 123 which did not increase upon cofactor supplementation (**Extended data Fig. 2b, c**), while those of 60 individuals had MMUT activity similar to controls (other MMA) (**Fig. 1b**).

**Fig. 1:**
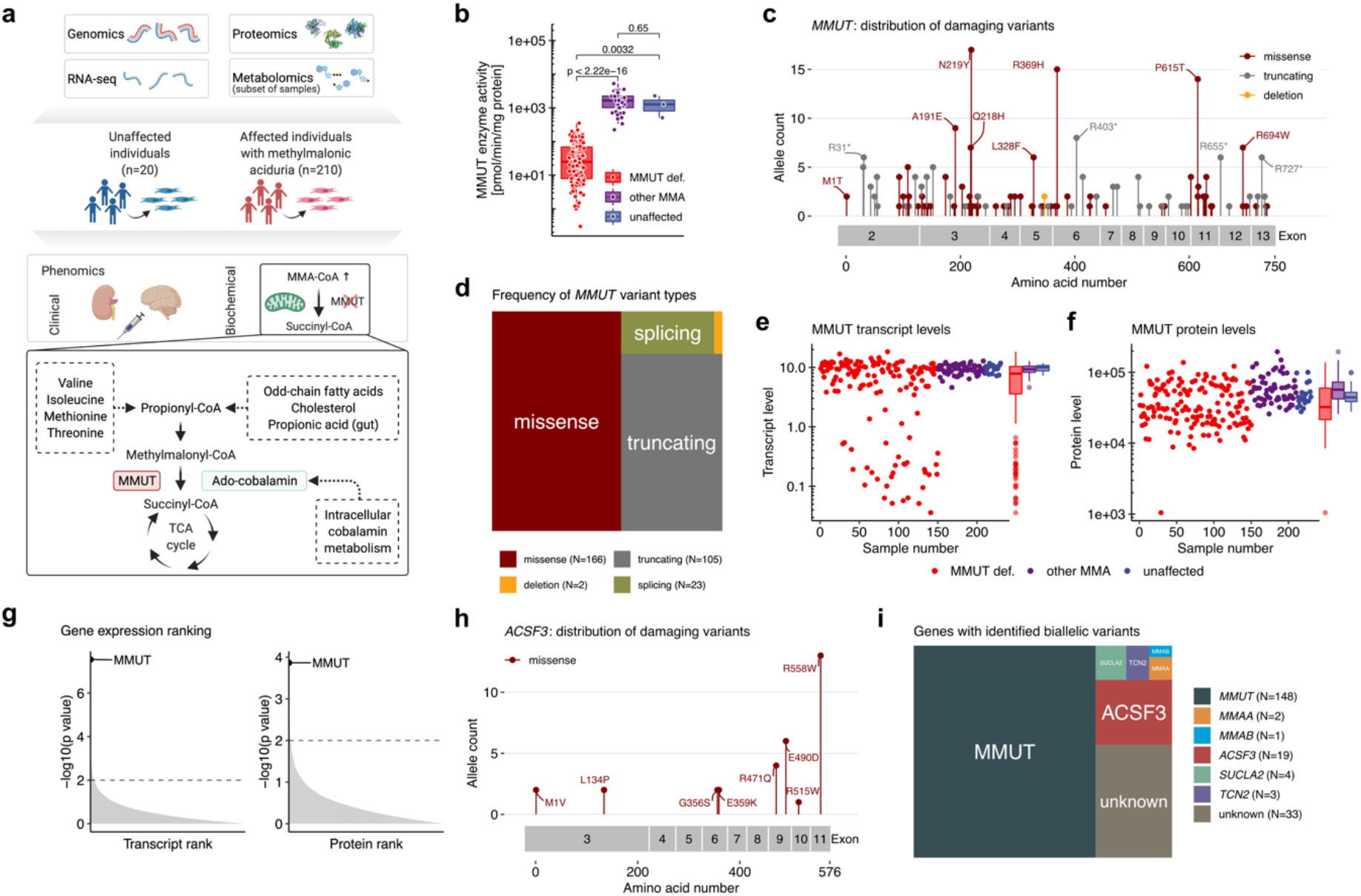
Multi-facetted omics view enabled a molecular diagnosis in 84% of individuals. **a**, Study overview with depiction of the propionate pathway including its precursors and the pathways catalyzed by MMUT. **b**, MMUT enzyme activity per study sub cohort. **c**, Lollipop plot of all pathogenic variants found on the *MMUT* gene. **d**, Proportions of variant types as identified on the *MMUT* gene. **e**, Transcript levels of *MMUT* by study sub cohorts. **f**, MMUT protein levels divided in study sub cohorts. **g**, Gene ranks according to p-values as calculated by gene-wise Welch’s *t*-test in the proteomics and transcriptomics data. **h**, Lollipop plot of pathogenic variants identified in *ACSF3*. **i**, Proportions of affected genes identified in the whole cohort.

In the MMUT-deficient samples we searched the WGS dataset for disease-causing variations in the *MMUT* gene and identified the molecular cause of disease in 148/150 individuals (**Extended data Table 1**). Pathogenic variants constituted 165 missense alleles, 105 truncating alleles, 21 splicing alleles, two alleles with in-frame deletions and three alleles containing copy number variants (**Fig. 1c, d** and **Extended data Fig. 2d**), of which 41 variants were novel (**Extended data Table 1**). RNA-seq identified reduced *MMUT* RNA expression in cells from MMUT-deficient individuals compared to the other groups (**Fig. 1e**). Individuals with strongly reduced RNA expression were enriched for splicing and/or truncating variants, consistent with nonsense-mediated decay (**Extended data Fig. 2e**). DIA-MS based proteome measurements revealed reduced MMUT protein levels in MMUT-deficient primary fibroblasts (**Fig. 1f**) which was distributed across all variant types (**Extended data Fig. 2e**). Consistent with its disease-causing role, MMUT RNA and protein levels were positively and significantly associated with PI and MMUT activity (**Extended data Fig. 2f**) and MMUT represented the most significantly dysregulated RNA and protein of MMUT-deficient samples when compared against all other samples (**Fig. 1g** and **Extended data Fig 2g**).

In the remaining 60 samples, we identified bi-allelic disease-causing variants for 22 individuals in genes other than MMUT: *ACSF3* (17 individuals) (**Fig. 1h**), *TCN2* (3 individuals), *SUCLA2* (1 individual), and MMAB (1 individual) according to the ACMG classification (**Extended data Table 1**). By searching RNA-seq for aberrantly expressed genes using OUTRIDER ^19^ (**Extended data Fig 3**), we identified two individuals with very low *ACSF3* expression; two with aberrant *SUCLA2* expression, in whom we confirmed predicted splicing and copy number variants at the genomic level (**Extended data Table 1**); two with very low *MMAA* expression, confirmed by complementation analysis; and one with low *MMAB* transcript, also confirmed by complementation analysis. Therefore, we identified a molecular cause for 29/60 remaining individuals (48%), including 18 pathogenic mutations, of which 10 were novel (**Extended data Table 1**). In sum, we found a diagnosis for 177/210 (84%) affected individuals (**Fig. 1i**), including 150 with deficiency of MMUT and 19 with damaging variants in *ACSF3* accounting for the largest cohort of ACSF3 deficiency.

### Phenotypic description and association to disease severity

It was our expectation that the genetic underpinnings identified above would only partly predict the clinical and biochemical phenotypes of affected individuals. We therefore aimed at establishing a quantitative assessment of disease severity by converting the catalogue of mostly semantic phenotypic traits into key numeric variables. A correlation matrix of all phenotypic variables (*n* = 105), spanning clinical symptoms at presentation and during disease course (*n* = 53), clinical treatments and therapeutic response (*n* = 23), clinical chemistry of blood or tissues including metabolite measurements (*n* = 13), and *in vitro* biochemical parameters (*n* = 13), revealed a cluster of features (MMUT activity, PI) that showed strong correlation across many variables (**Fig. 2a, b**).

**Fig. 2:**
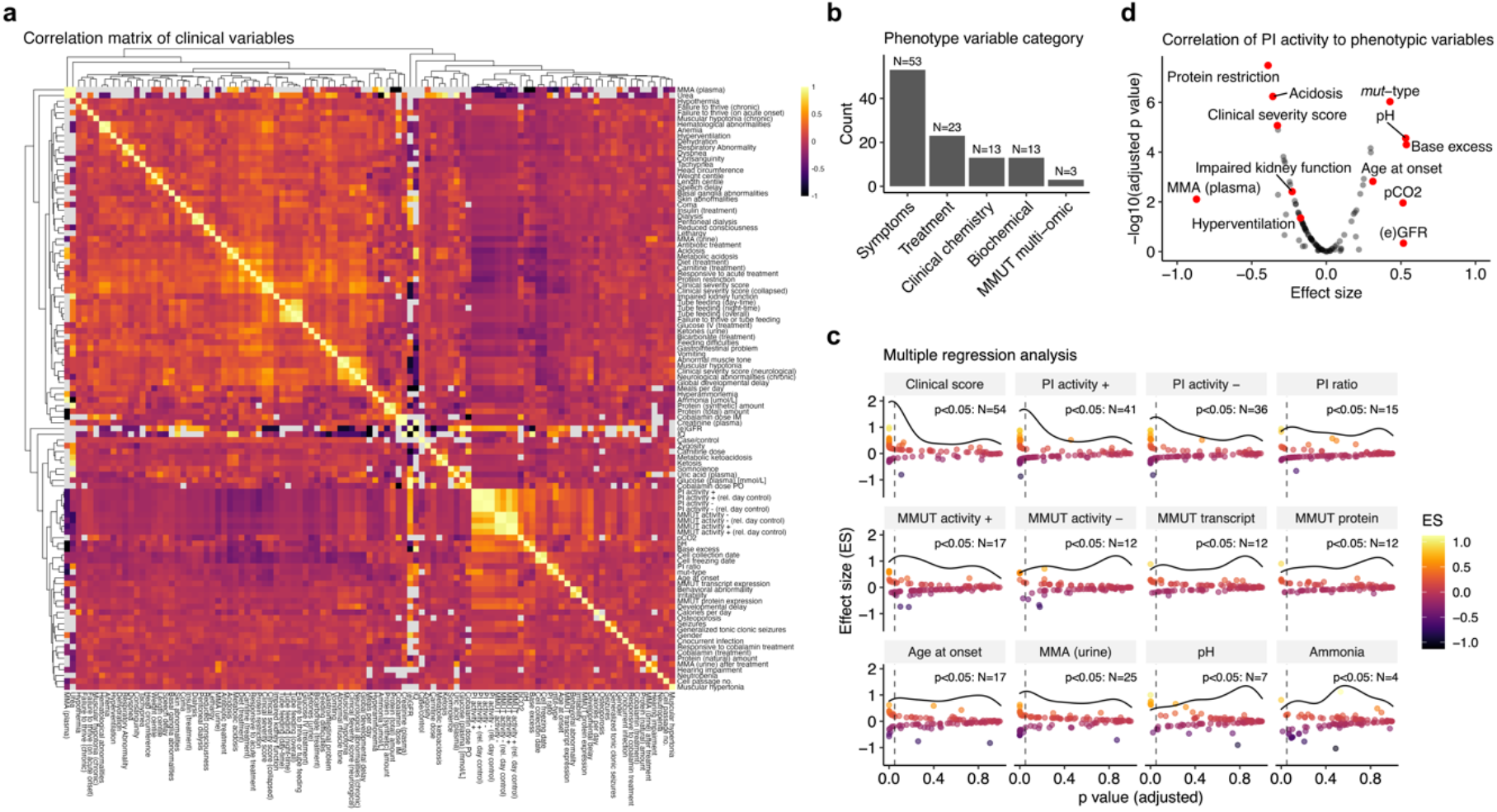
Phenomics analysis reveals two main surrogate markers of disease severity (clinical severity score and PI+ activity). **a**, Correlation matrix of all continuous numeric and discrete phenotype variables. **b**, Number of phenotypic traits according to five phenotype subcategories. **c**, Panel of selected phenotypic traits and their overall strength of representing the entirety of the phenomics dataset (here termed clinical disease severity) as assessed by linear modelling. Each point represents the result of linear regression against one other phenotypic variable with the effect size on the y-axis and the resulting p-value on the x-axis. The horizontal curved line indicates the density of data points as distributed along the x-axis. Vertical dashed line indicates threshold of significance (p-value <0.05). **d**, Linear regression results of the PI+ activity variable compared against the rest of the phenotypic variables. Phenotypic traits available from **Source data Table 1**.

Since the identified few variables strongly associated with many clinical features, we postulated that most disease characteristics might be well predicted by one or a select few variables. As proof-of-principle, we established a clinical severity score (CSS), which incorporated the outcome of five typical clinical features ^20^ (for composition see **Methods**), whereby a score of 0 represented absence of these typical MMA features, a score of 1 mild MMA, and a score of 2 or higher (max. 5) moderate to severe MMA disease. Comparison of the CSS against all phenotypic parameters demonstrated significant correlation with 54 individual variables (**Fig. 2c**), including many classical phenotypic symptoms of MMAuria, such as acidosis, hyperammonemia and muscular hypotonia as well as the requirement for dietary and pharmacological interventions (**Extended data Fig. 4a**). Importantly, the CSS also inversely correlated with age of onset (**Extended data Fig. 4b**), a parameter that on its own has been used as an indication of clinical severity ^21^.

Multiple correlation analysis identified PI in the presence of hydroxocobalamin (PI+) to significantly correlate to 41 phenotypic features, the most of any individual continuous variable (**Fig. 2c**). This contrasts, for example, to age at onset which significantly correlated with only 17 parameters (**Fig. 2c**). Closer inspection revealed PI+ to be inversely correlated to disease severity, including significant positive correlation with, e.g., glomerular filtration rate or age at disease onset, and negative correlation with, e.g., methylmalonic acid concentration in plasma, presence of clinical interventions such as protein restriction and the CSS (**Fig. 2d** and **Extended data Fig. 4c, d)**. Therefore, in line with its validity as a diagnostic test for MMA ^18^, the PI+ variable was used as an approximation of clinical disease severity in this study.

### Multi-layered biology reveals disruption of TCA cycle and associated pathways

To identify disease-associated expression alterations of genes, proteins, and pathways, we attempted a global assessment of transcript and protein expression, integrated with the quantitative phenotype variables identified above. Since patients with TCN2, SUCLA2 and ACSF3 deficiency lack most of the typical signs and symptoms of classical MMA, we compared MMUT-deficient with all non-MMUT-deficient samples (control).

Investigation of transcripts and proteins using differential correlation patterns (Pearson correlation method), dimensionality reduction via Principal Component Analysis and DESeq2 did not immediately yield clear grouping of the data, nor obvious expression pattern differences between the groups (**Extended data Fig. 5a, b, c**). However, multi-omics factor analysis ^22^, integrating both genetic data layers and proteotyping data, identified mitochondrial metabolic pathways and in particular the electron transport chain and the TCA cycle to be enriched in MMUT-deficient samples (**Fig. 3a**). In more detail, the proteins SLC16A3, CS, MDH2, and OGDH were found to be the main drivers of this particular factor’s variance in the proteotyping data within the TCA-associated gene sets (**Fig. 3b**). Linear discriminant analysis of genes shared between transcriptomics and proteotyping indicated MMUT as the strongest and SUCLA2, OGDH, and PDHB to be top drivers of separation between MMUT-deficient and control samples (**Fig. 3c**). Further, gene set enrichment analysis utilizing sample stratification by disease severity, both by CSS and PI+, also identified oxidative phosphorylation and the TCA cycle as over-represented pathways in the proteomics (CSS and PI+) and transcriptomics (CSS) datasets (**Fig. 3d**).

**Fig. 3:**
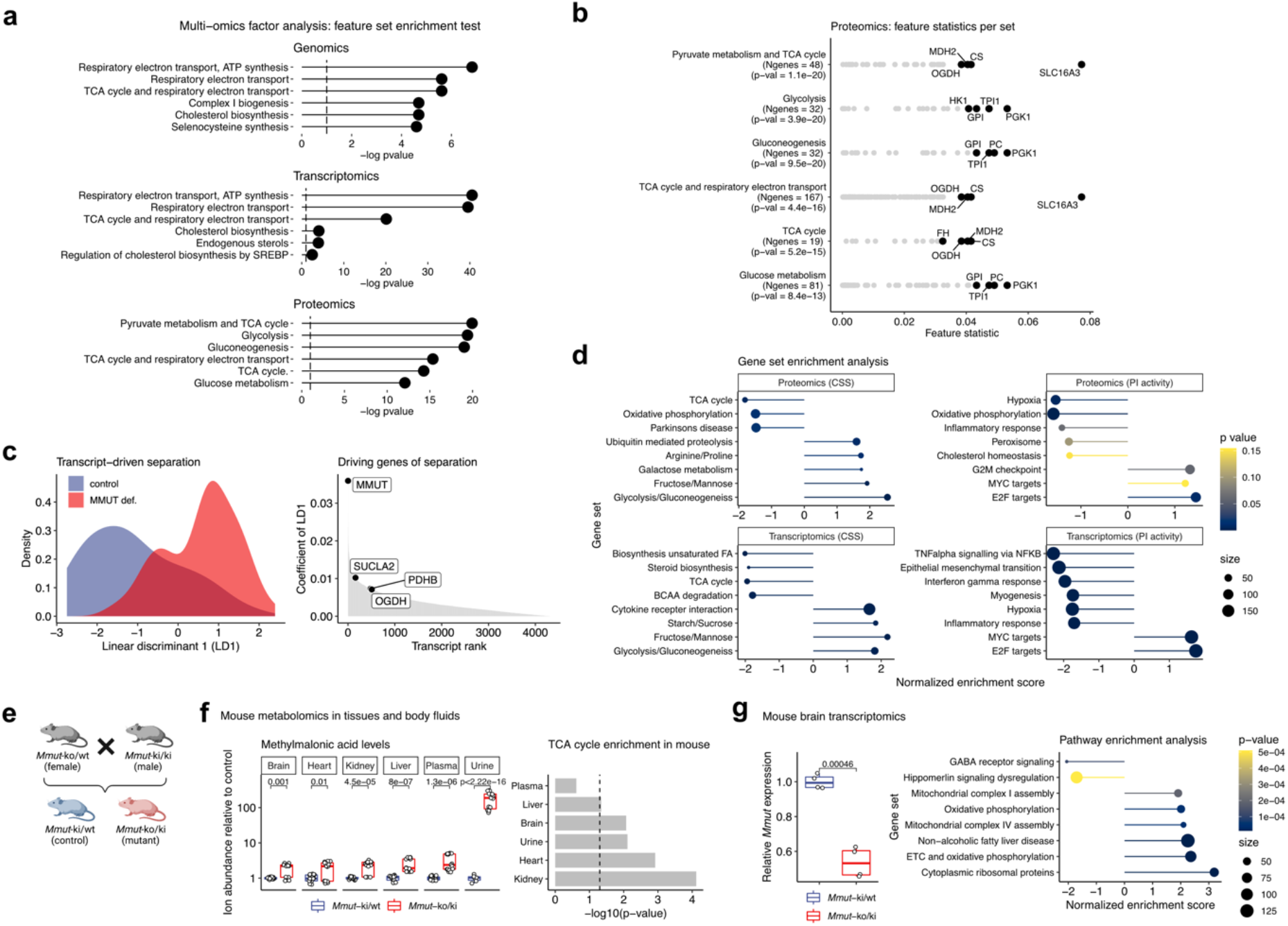
Untargeted integration of omics data layers highlights the TCA cycle and associated pathways as well as oxidative phosphorylation gene sets to be dysregulated in MMA. **a**, Gene set enrichment test using the multi-omics factor analysis tool (MOFA). **b**, Detailed feature statistics of the top enriched gene sets following MOFA in the proteomics data. **c**, Linear discriminant model (split to assign training and test data: 0.5) of transcripts separates MMUT-deficient from control driven by *MMUT* and other genes related to the TCA cycle. **d**, Gene set enrichment analysis based on effect size ranking derived from differential expression analysis (also see **Fig 4b**). **e**, Breeding scheme of *Mmut* deficient mice. **f**, Untargeted metabolomics in mouse tissues and body fluids, depicting boxplots for methylmalonic acid and metabolite set enrichment analysis based on the complete metabolomics dataset. **g**, RNA-seq on mouse brain tissue. Boxplots of the relative *Mmut* transcript abundance and gene set enrichment analysis following DESeq2 analysis, dot size represents number of genes per set.

The biological relevance of these changes was confirmed in a hemizygous mouse model of MMA ^23^ (**Fig. 3e**). Untargeted metabolomics of brain, heart, kidney, liver, plasma, and urine confirmed elevated levels of the eponymous metabolite methylmalonic acid in mutant animals, while pathway enrichment analysis pointed to dysregulated TCA cycle pathways in all tissues and urine (**Fig. 3f**). Transcriptomics of brain tissue further confirmed the expected 50% reduction in *Mmut* transcript of mutant mice, along with enrichment of electron transport chain and oxidative phosphorylation pathways (**Fig. 3g** and **Extended data Fig. 6**).

### MMUT deficiency leads to transcript and protein alterations in proximal enzymes

Since both data-driven and phenotypically stratified analyses indicated TCA and associated pathways to be disrupted in disease, we performed a concerted investigation of the TCA cycle enzymes, including those which metabolize anaplerotic (replenishing TCA cycle intermediates) and cataplerotic (removing TCA cycle intermediates) reactions, from which we had both RNA and protein information. As controls we included isoforms of TCA enzymes which are not involved in these pathways (**Fig. 4a**). Direct comparison of RNA and protein expression between MMUT-deficient and control cells revealed MMUT to be significantly dysregulated (**Fig. 4a outer band**).

**Fig. 4:**
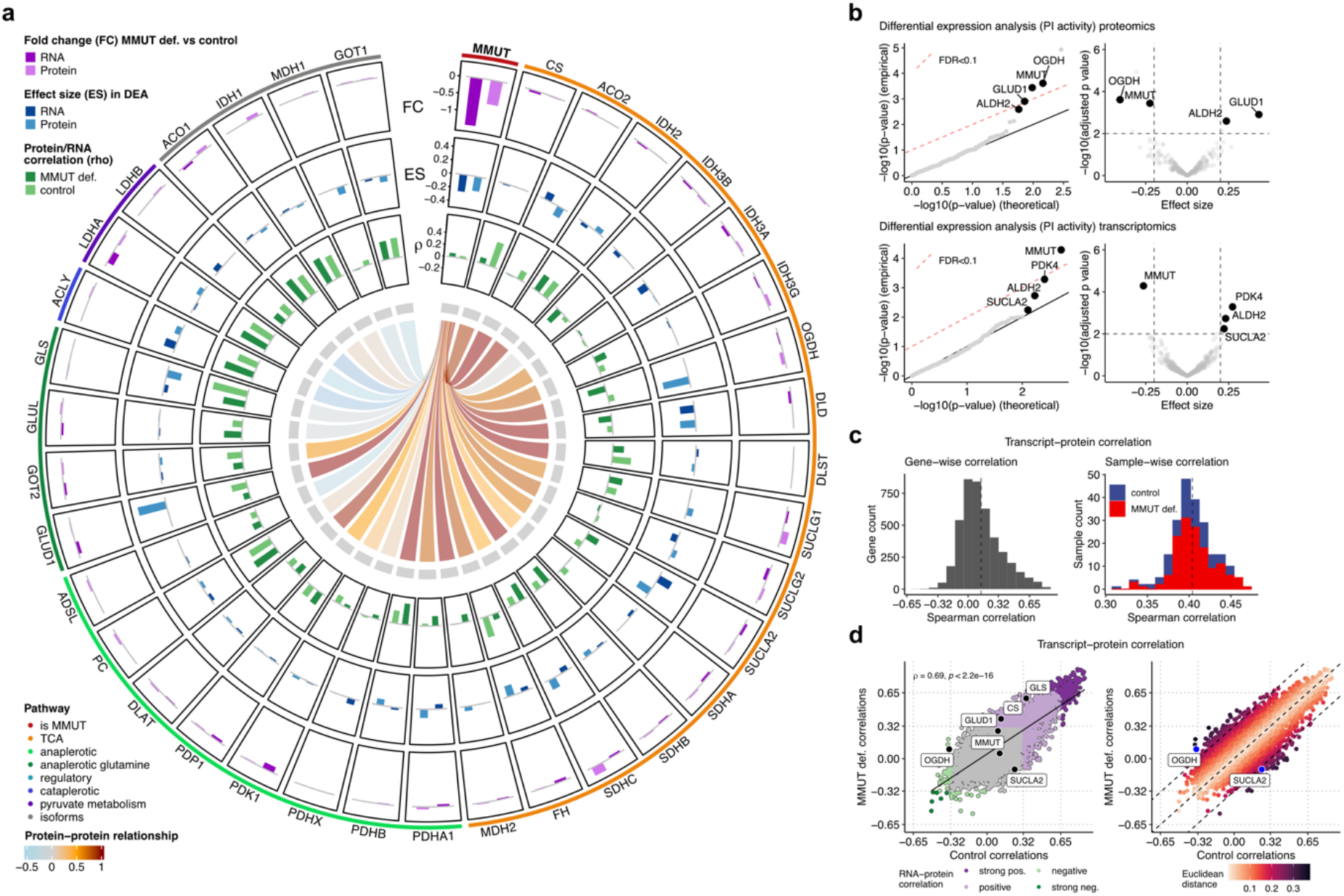
Transcript-protein and protein-protein correlation analyses reveal coordinated relationships between MMUT and TCA genes and proteins but not their isoforms. **a**, Circos plot depicting raw fold changes of transcripts and proteins, effects sizes derived from differential expression analysis, transcript-protein correlations, and correlative relationships of the MMUT protein to TCA proteins and their corresponding isoforms. **b**, Q-Q and volcano plots illustrate the results of the differential expression analysis based on a linear mixed modeling approach applied to the proteomics and transcriptomics data, restricted to enzymes (or their encoding genes) localized in the mitochondria. **c**, Histograms of Spearman correlations across 4318 transcript-protein pairs (left), and 221 samples (right). **d**, Scatter plot of Spearman correlations in MMUT-deficient against control. Euclidean distance from the diagonal is calculated based on the formula |(MMUT def. correlation – control correlation)|/sqrt(2).

Differential expression analysis, performed using a linear mixed modelling approach ^24^, identified the genes with the strongest effect size and significance to be enriched for mitochondrial localization, as listed in MitoCarta 3.0 ^25^ (**Extended data Fig. 7a**). Closer examination (**Fig. 4a middle band, b**), identified MMUT to be significantly downregulated in disease at both the RNA and protein level, while ALDH2, which catalyzes the interchange between methylmalonate and methylmalonate semialdehyde, was in contrast upregulated in both. A further upregulated transcript was *PDK4* (**Fig. 4b**), which is responsible for the phosphorylation and as a consequence inactivation of the pyruvate dehydrogenase complex. However, the proteins with the overall largest effect size were OGDH (downregulated in disease) and GLUD1 (upregulated in disease), both enzymes involved in the anaplerosis of glutamine (**Fig. 4b**).

Examination of RNA-protein expression correlation in all samples revealed a limited Spearman correlation of 0.14 at the gene level (4318 transcript-protein pairs) and 0.40 at the sample level (**Fig. 4c** and **Extended data Fig. 8a**), similar to findings by others ^26^. Comparison of RNA-protein correlation in MMUT-deficient cells compared to controls revealed that, while 1158 pairs (26.8%) correlated significantly (p-value < 0.05) in both genotypes (**Fig. 4d** all colored points and **Extended data Fig. 8b**) in accordance with previous studies ^27^, the correlation of some genes segregated depending on the genotype (MMUT-deficient vs control) (**Fig. 4d**). In particular, OGDH, GLUD1, CS, and GLS showed higher RNA-protein correlation in MMUT-deficient samples than controls, while SUCLA2 had reduced RNA-protein correlation (**Fig. 4a, d** and **Extended data Fig. 8c**). OGDH and SUCLA2 were among the genes with the strongest genotype-dependent RNA-protein correlation changes (**Fig. 4d**). Interestingly, we found poor RNA-protein correlation for MMUT in both control and MMUT-deficient cells (**Fig. 4a, d** and **Extended data Fig. 8c**).

Finally, MMUT protein levels positively correlated to protein levels of many TCA and anaplerotic enzymes in control but not in MMUT-deficient cells, while there was little or no protein expression correlation between MMUT and non-TCA protein isoforms in either genotype (**Fig. 4a center** and **Extended data Fig. 8d**). Such a relationship is exemplified by MMUT:ACO2 and MMUT:ACO1 (**Fig. 4a center** and **Extended data Fig. 8e**) and provides the first indication that MMUT may be part of a so far unknown interaction network with these mitochondrial TCA cycle and anaplerotic enzymes^28^. Examination of pairwise correlation between all proteins and transcripts (**Extended data Fig. 8f, g**) in these pathways suggests that TCA cycle and anaplerotic enzymes have a positive correlation with each other, which is not altered in MMUT deficiency, unless MMUT is included in the comparison. Overall, the above findings suggest that disruption of MMUT RNA and protein expression drives regulatory changes in certain TCA and anaplerotic enzymes.

### Metabolomics highlights rewiring of TCA cycle anaplerosis

To examine the functional consequences of the above RNA and protein expression alterations, we performed untargeted metabolomic analysis on a set of 6 MMUT-deficient and 6 control primary fibroblasts derived from unaffected individuals (for selection criteria, see **Extended data Fig. 9a** and **Methods**). Among the most significantly changed metabolites we found decreased glutamine and alanine as well as increased hexoses, methylcitrate, oxoadipate, aminoadipate, and pyruvate (**Fig. 5a**). Combining these alterations with observed changes in RNA and protein expression in the same samples provides insight into disruption of TCA cycle anaplerosis in MMUT deficiency (**Fig. 5b**).

**Fig. 5:**
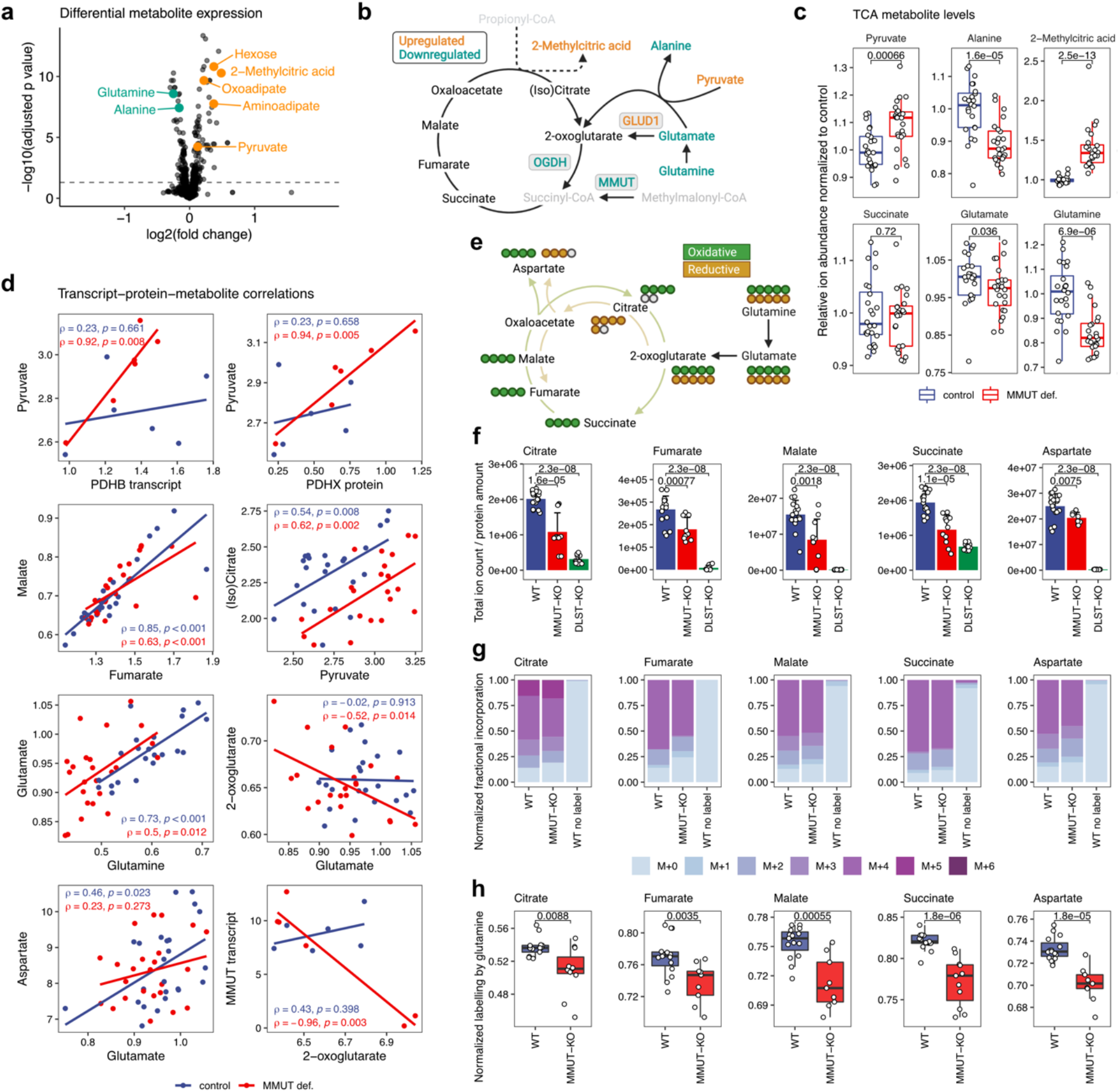
Polar metabolomics in selected patient fibroblasts and glutamine tracing studies in CRISPR/Cas9 KO 293T cells highlight differential glutamine anaplerosis. **a**, Volcano plot depicting differentially expressed metabolites. Highlighted are the ones particularly relevant to this study. **b**, Schematic depiction of the TCA cycle and relevant anaplerotic reactions. The color code indicates dysregulations on metabolite and protein level; grey metabolites were not detected. **c**, Boxplots to compare relevant TCA cycle metabolites, including the pathologically relevant biomarker 2-methylcitrate formed upon condensation of oxaloacetate with propionyl-CoA. **d**, Pearson correlation of a certain key metabolite against a metabolite, transcript, or protein variable. **e**, Schematic representation of labeling of TCA cycle and associated metabolites derived from labelled glutamine via anaplerosis. Circles represent carbon atoms. **f**, Pool sizes of metabolites in control and CRISPR/Cas9 KO 293T cells. **g**, Relative abundance of isotopologues after glutamine labelling. **h**, Relative fractional incorporation of labelled glutamine into TCA cycle and associated metabolites. All p-values are calculated by Wilcoxon rank test.

The increased pyruvate level (**Fig. 5a, c**), increased expression of the PDH complex inhibitor *PDK4* (**Fig. 4b**), and strongly elevated 2-methylcitrate (**Fig. 5c**), a biomarker of MMA formed by citrate synthase mediated mis-condensation of oxaloacetate with propionyl-CoA instead of acetyl-CoA, together suggest reduced incorporation of pyruvate into the TCA cycle. This is further supported by the analysis of our cross-dimensional data which identified stronger pyruvate to *PDHB* transcript and pyruvate to PDHX protein correlations (**Fig. 5d**), implying reduced pyruvate turnover per transcript and protein respectively. Consecutive metabolites in a pathway often correlate strongly ^27^, here exemplified by malate and fumarate (**Fig. 5d**). Although we found such a correlation between pyruvate and its downstream metabolite (iso)citrate in both control and MMUT-deficient cells, our data suggests that in MMUT-deficient cells a higher concentration of pyruvate is required to create the same amount of (iso)citrate as controls (**Fig. 5d**), again supporting reduced incorporation of pyruvate into the TCA cycle.

Despite this reduced pyruvate-mediated anaplerosis, 2-oxoglutarate, succinate, fumarate and malate levels remained unchanged in MMUT-deficient cells (**Fig. 5c** and **Extended data Fig. 9b**), suggesting stable TCA cycle metabolite pools in these primary fibroblast cells. As our previous findings indicated altered proteins in glutamine anaplerosis (**Fig. 3b, c, 4a**), we investigated the contribution of this pathway towards the balance of the TCA cycle. Glutamine strongly correlated with glutamate in both MMUT-deficient and control samples (**Fig. 5d**). However, in MMUT-deficient cells, glutamine and glutamate were decreased (**Fig. 5c**), and there was a negative correlation of glutamate with 2-oxoglutarate, suggestive of more tightly regulated glutamine flux under disease conditions (**Fig. 5d**). Abrogated positive correlation of glutamate and aspartate potentially points to an altered dependence on glutamate for TCA cycling but not production of cataplerotic metabolites, such as aspartate (**Fig. 5d**). In line with this, we found an inverse relationship between *MMUT* transcript and 2-oxoglutarate in MMUT-deficient cells (**Fig. 5d**). While we focused our efforts on the investigation of TCA anaplerosis at this stage, the power of our untargeted dataset was further illustrated by more unexpected findings, including increased oxoadipate and aminoadipate (**Fig. 5a**), upstream metabolites of 2-oxoadipate dehydrogenase complex which shares its E2 (DLST) and E3 (DLD) components with 2-oxoglutarate dehydrogenase complex (**Extended data Fig. 9c**), suggestive of a preference for 2-oxoglutarate over 2-oxoadipate metabolism.

The above results point to an adjusted reliance on the glutamine anaplerotic pathway in disease; a hypothesis we tested further by assessing relative carbon incorporation derived from anaplerotic glutamine into TCA cycle and associated intermediates using MS-based stable isotope tracing. For this experiment, control (wildtype, WT), *DLST*-KO, and *MMUT*-KO 293T cells, validated by Western blotting and enzyme activity measurements (**Extended data Fig. 10**), were cultured in glutamine-free glucose-DMEM supplemented with [U-^13^C]glutamine. *DLST*-KO cells were used as a control to mimic the above detected reduction of OGDH protein in MMUT deficiency, allowing us to assess fractional incorporation of glutamine into TCA cycle intermediates when the oxidative pathway (clockwise from 2-oxoglutarate, **Fig. 5e**) is completely blocked (DLST is a component of the 2-oxoglutarate dehydrogenase complex, metabolizing 2-oxoglutarate to succinyl-CoA). By analyzing the pool sizes, we could confirm reduced levels of glutamine and glutamate (**Extended data Fig. 11a**) comparable to the patient fibroblasts (**Fig. 5c**), validating our experimental model. In addition, we found markedly reduced TCA cycle metabolites, including citrate, fumarate, malate, succinate, and aspartate (a proxy for oxaloacetate ^29^), indicating an overall reduced TCA metabolite pool in MMUT deficiency, while KO of *DLST* led to virtual absence of most TCA cycle metabolites (**Fig. 5f**). To identify differential labeling patterns, we studied the isotope distribution based on the relative incorporation of glutamine-derived carbons and found a decreased proportional fraction of M+4 isotopologues under *MMUT*-KO conditions for all studied TCA cycle metabolites and aspartate (**Fig. 5g, Extended data Fig. 11b**), suggesting reduced derivation from glutamine. Indeed, we found a decreased fractional contribution of glutamine to the TCA cycle for all studied intermediates (**Fig. 5h**). Moreover, consistent with reduced OGDH activity, there was a relative preference for the reductive TCA cycle pathway, as indicated by an increased M+5/M+4 ratio for citrate ^30,31^ in MMUT deficiency (**Extended data Fig. 11c**). In sum, this suggests there is reduced production of TCA cycle intermediates which is not compensated for by glutamine anaplerosis.

### MMUT physically interacts with 2-oxoglutarate dehydrogenase and other anaplerotic enzymes

The above observed adjustments in glutamine anaplerosis have an unclear regulatory etiology. Based on the strong protein expression correlation between MMUT and proximal TCA cycle and anaplerotic enzymes (**Fig. 4a** and **Extended data Fig. 8d)**, we hypothesized that these proteins may be part of a shared metabolon complex, potentially facilitating regulation of TCA cycle anaplerosis of glutamine by protein-protein interactions ^28^. To gain insights into this potential physical relationship, we took advantage of over-expression of C-terminally flag-tagged versions of MMUT, two pathway members (MMAB and MCEE) expected to participate in any multi-protein complex containing MMUT, and two negative controls (VLCAD and empty vector) in 293T cells (**Fig. 6a**). Using cross-linking affinity purification coupled to mass spectrometry, at 1.0% FDR using 2 peptides minimum at 95% threshold, each of these ‘bait’ proteins pulled-down a total of 100-350 different ‘prey’ proteins over 3 biological replicates (**Fig. 6b**). Within this intersection, we identified 57 prey proteins pulled down by MMUT, MMAB and MCEE in at least one replicate, but not by empty vector or VLCAD in any replicate (**Fig. 6b**).

**Figure 6.**
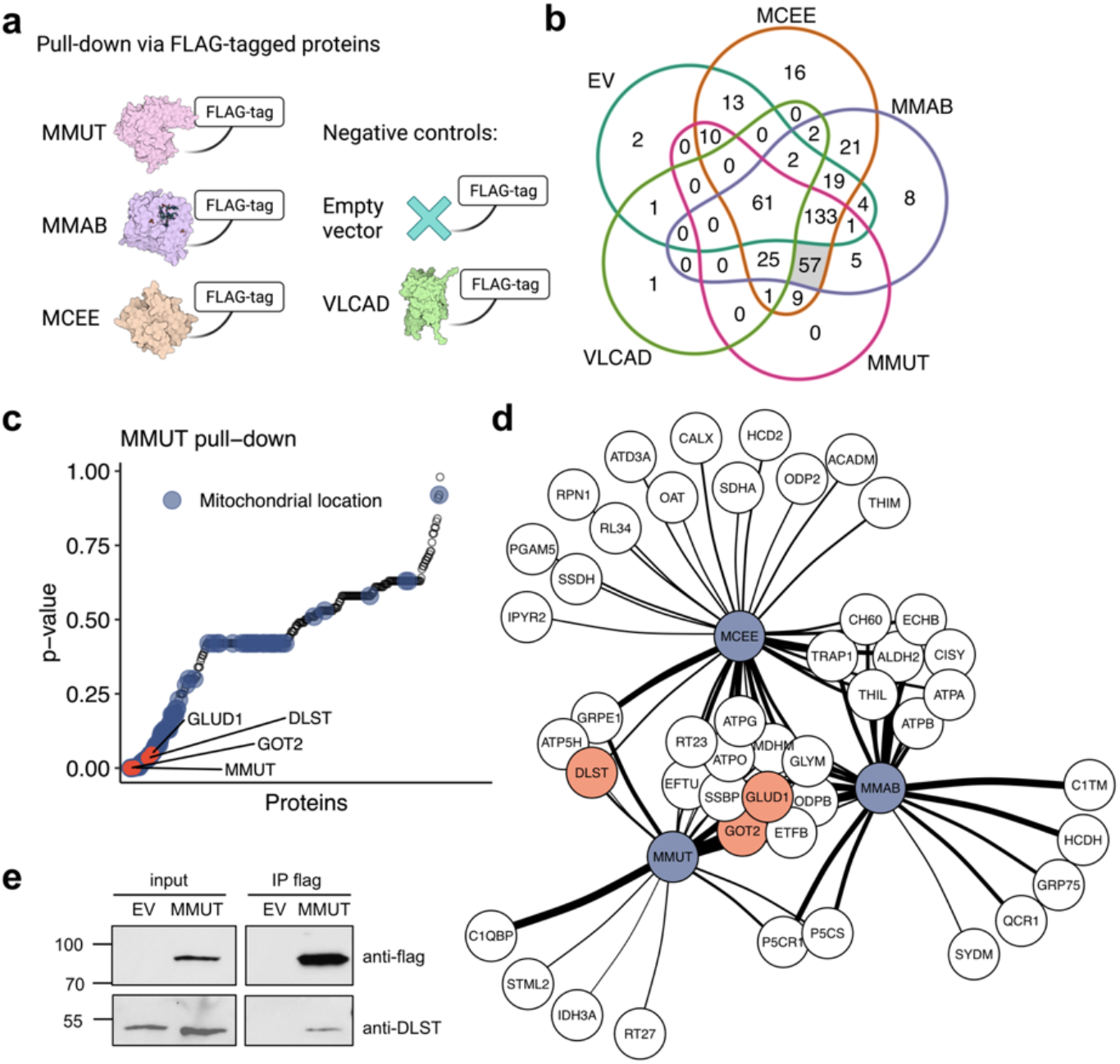
MMUT interacts physically with GLUD1, DLST, and GOT2 as demonstrated by FLAG-tag pull-down. **a**, Outline of experimental and control groups indicating which protein was used with a FLAG-tag in a cross-linking affinity purification experiment coupled to subsequent analysis of the pull-down samples by mass spectrometry. **b**, Venn diagram of proteins pulled down by the different flag-tagged proteins. **c**, ANOVA p-values of all proteins pulled down by MMUT. Blue dots indicate proteins with mitochondrial location according to Uniprot. **d**, Interaction network of significantly enriched proteins (ANOVA p-value < 0.05). Thicker connector indicates lower p-value. **e**, Western blot of immunoprecipitation of flag-tagged MMUT probing for DLST.

Analysis of variance (ANOVA) of the biological triplicates comparing MMUT with EV and VLCAD identified 22 proteins to be significantly enriched (nominal p-value <0.05) in the MMUT sample (**Fig. 6c** and **Extended data Table 2**). All proteins were designated by Uniprot to have mitochondrial localization and included GLUD1, GOT2 and DLST (**Fig. 6c**). ANOVA comparing the intersection of proteins confidently pulled down by all three of MMUT, MMAB and MCEE but not of EV or VLCACD identified 11 MMUT/MMAB/MCEE interacting proteins, including GLUD1 and GOT2; while the intersection of two of MMUT, MMAB, and MCEE against both negative controls identified 13 proteins which interacted with MMUT/MMAB, MMUT/MCEE or MMAB/MCEE, including DLST (**Fig. 6d** and **Extended data Table 2**). Interaction of MMUT and DLST was further confirmed by Western blot analysis (**Fig. 6e** and **Extended data Fig. 12ab**). These data indicate that MMUT physically interacts with GLUD1 and the oxoglutarate dehydrogenase complex component DLST and suggests that disruption of this interaction may underlie their altered regulation in disease.

## Discussion

In this study, we used an integrated multi-modal approach to diagnose and uncover new pathomechanisms of the inborn error of metabolism MMA. Unique to this investigation was the large set of patient samples along with corresponding phenotypes available, and the ability to coordinate aliquots from the same samples to generate data at three molecular layers. The results of our study will encourage future endeavors to use our approach in any setting of an inborn monogenic disease. Moving forwards, the datasets derived from our study can be further exploited, e.g., by applying network contextualization tools ^32^, integrating multi-omics and flux modeling ^33^, and reconstructing genome-scale metabolic networks ^34^, continuing to refine the pipeline of a multi-modal study of inborn errors of metabolism.

Our findings reinforce the value of comprehensive and complementary datasets to increase diagnostic yield and the understanding of the pathophysiological underpinnings of disease. Our multi-modal profiling allowed the identification of causative genetic variation in 84% of the cohort, including causative factors in the samples without MMUT deficiency. We were able to widen the set of genes beyond the classical MMA genes *MMUT, MMAA*, and *MMAB*. For example, the identification of *ACSF3* damaging variants in our cohort is particularly interesting as they have recently been linked to combined malonic and methylmalonic aciduria (CMAMMA) ^35^. The phenotype of CMAMMA patients was indistinguishable from the remainder of MMA patients with normal MMUT activity, highlighting the fact that inborn errors of metabolism present with widely overlapping phenotypes and that they should be studied with large gene panels or with WGS approaches to avoid biases towards known genes and to augment the chances of diagnosis.

While the ability of clinical phenotypic information to predict a molecular diagnosis was limited, phenotypic variables – both clinical and biochemical – enabled sample stratification by disease severity and consequently identification of multi-level alterations of metabolic genes/proteins which were not apparent following examination of single omics layers. Such a move away from “data silos” into true integrative and mechanism-based multi-layered analysis remains challenging, as it requires new analytical and statistical methods to combine these disparate data sets ^36^. In this capacity, multi-omics factor analysis ^22^ highlighted the disruption to transcripts and proteins of the TCA cycle and related pathways, a finding verified by the correlation with phenotypic data utilizing both propionate incorporation activity and a clinical severity score. Following multi-modal integration, we performed metabolomics in select patient cells and further complemented the data with glutamine tracing and protein-protein interaction studies in a second cell model. In sum, these experiments showed decreased TCA metabolite pools and a reduced glutamine-derived anaplerosis, in line with the detected reduction of OGDH. In addition, we identified a list of novel MMUT-interaction candidates, among which DLST (OGDH complex component) and GLUD1 are directly involved in the glutamine anaplerotic pathway. Abrogation of the interaction with MMUT, due to *MMUT* knockout, could explain the reduction of glutamine anaplerosis. It is of note, that such a tailored set of follow-up experimental approaches – orthogonal to multi-omics data – is invaluable for molecular assessment of potential targets and the validation of their biological significance.

Overall, our results highlight the importance of the loss of methylmalonyl-CoA as an anaplerotic source and indicate a relevant reduction of TCA cycle intermediates in MMA, in part due to reduced anaplerotic supply stemming from glutamine. For the first time we show anaplerotic insufficiency in MMA and propose this phenomenon as a novel therapeutic target. Anaplerotic stimulating approaches have precedent in IEMs, including application of triheptanoin in long-chain fatty acid oxidation disorders ^37^. Unfortunately, since fatty acid β-oxidation produces two acetyl-CoA and one propionyl-CoA per heptanoate molecule, use of this particular compound in MMUT deficiency is not feasible. However, anaplerotic treatment of patients suffering from the MMA-related disorder propionic aciduria with the TCA cycle intermediate citrate has been attempted, with inconsistent results ^38^. Based on our findings, compounds such as dimethyl oxoglutarate, a membrane-permeable alternative to 2-oxoglutarate, previously used in a model of OXPHOS dysfunction ^30^, may represent a more promising therapeutic strategy. Studies to delineate the efficacy of such approaches in preclinical and clinical models will be important for the ongoing development of new treatments for MMA and inborn errors of metabolism in general.

## Data Availability

All data produced in the present study are available upon reasonable request to the authors.

## Acknowledgements

This project was funded by the ETH domain strategic focus area “Personalized Health and Related Technology” (PHRT; https://www.sfa-phrt.ch). This work received financial support from the Swiss National Science Foundation [31003A_175779] to M.R.B. and [310030_192505] to D.S.F. and from the University Research Priority Program of the University of Zurich (URPP) ITINERARE— Innovative Therapies in Rare Diseases. P.F. was supported by the Filling the Gap grant awarded by the Medical Faculty, University of Zurich, Switzerland. We acknowledge the Functional Genomics Center Zurich of the University of Zurich for RNA sequencing of the mouse brain tissues and bioinformatics support as well as for support with the glutamine isotope studies.

## Author Contributions

Conceptualization, I.X., R.A., G.R., E.D., B.W., M.R.B., D.S.F.; Methodology, P.F., X.B., D.L., W.S., T.P., M.F., L.S., C.H., R.J.M., S.C., S.G., A.v.D., K.H., P.P., N.Z.; Investigation/Data analysis, P.F., X.B., D.L., W.S., T.P., C.F., A.B., S.C., M.P., S.G., P.P., F.T., N.Z., G.R., D.S.F.; Writing – Original Draft, P.F., D.S.F.; Writing – Review & Editing, all co-authors; Visualization, P.F., X.B., D.L., W.S., C.F.; Supervision, P.F., K.H., P.P., F.T., I.X., G.R., E.D., B.W., D.S.F.; Project Administration, P.F., B.W.; Funding Acquisition, R.A., B.W., M.R.B., D.S.F.

## Extended data

Supplementary figures and tables in separate files.

## Source data

### Source data Proteomics

All mass spectrometry proteomics raw files have been deposited to the ProteomeXchange Consortium via the MassIVE partner repository (https://massive.ucsd.edu) with the dataset identifier MSV000088791.

### Source data Metabolomics

Metabolomics mass spectrometry raw data for human fibroblast measurements have been uploaded to the MassIVE data repository (https://massive.ucsd.edu) with the dataset identifier MSV000089082.

### Source data Affinity capture mass spectrometry raw data

IP-MS raw files have been deposited to the ProteomeXchange Consortium via the MassIVE partner repository (https://massive.ucsd.edu) with the dataset identifier MSV000088791.

### Source data Quantitative Proteomics Data

CSV file containing the quantitative proteomics data used for the analyses.

### Source data Quantitative Transcriptomics Data

CSV file containing the quantitative RNA-seq data used for the analyses.

### Source data Table 1 Phenotypic dataset

CSV file containing all clinical information.

### Source data Table 2 Pull-down

Complete list of proteins that were pulled down in the affinity capture mass spectrometry experiment.

## Methods

### Cohort and patient-derived fibroblast samples

Primary fibroblast samples and corresponding disease-related information, including clinical and diagnostic data, was collected from 1989 to 2015. The information was obtained and used under the ethics approval granted by the Ethics Committee of the Canton of Zurich, Switzerland (no. KEK-2014-0211, amendment: PB_2020-00053). Upon collection, primary fibroblasts were cultured using Dulbecco’s modified Eagle medium (Gibco, Life Technologies, Zug, Switzerland) with 10% fetal bovine serum (Gibco) and antibiotics (GE Healthcare, Little Chalfont, UK) and either used immediately, or exchanged to 90% fetal bovine serum and 10% dimethyl sulfoxide and stored in cryovials under liquid nitrogen. A frozen aliquot of each primary fibroblast cell culture was sent for WGS, RNA-seq and DIA-MS analysis (**Fig. 1a**). RNA-seq and DIA-MS were always performed from matched aliquots.

### Clinical disease severity score

The clinical disease severity score was based on five typical clinical signs/symptoms of MMA ^20^, including age at disease onset, as well as the presence of neurological abnormalities, kidney impairment, hematological abnormalities, and failure to thrive. Each patient was assigned a score from 0-5 indicating increasing disease severity (**Source data Table 1**).

### Biochemical assays

Propionate incorporation into acid precipitable material of primary fibroblasts was assessed according to a protocol described previously ^39^ with modification as described ^18^. Methylmalonyl-CoA mutase enzyme activity assay was performed in crude cell lysates as originally described ^40,41^ using recent modifications ^14^.

### Whole genome sequencing

Genomic DNA was isolated using the QIAmp DNA Mini Kit reagents (Qiagen; Hilden, Germany) following the protocol provided by the supplier. DNA was quantified by Qubit (Thermo Fisher; Waltham, USA). Whole genome sequencing libraries were prepared with the TruSeq DNA PCR-free library reagents (Illumina; San Diego, USA) using 1ug of genomic DNA following the protocol provided by the supplier. The genomic DNA libraries were quantified using the KAPA Library Quantification Complete Kit (Roche; Basel, Switzerland) according to the protocol supplied with the reagents. The quantified libraries were sequenced on the NovaSeq 6000 sequencer (Illumina) using a 150-nucleotide paired end run configuration following the protocol provided by the supplier.

### RNA sequencing

Total RNA was isolated using the RNeasy Plus Mini Kit (Qiagen). The total RNA was quantified using the Qubit (Thermo Fisher) and quality-controlled using the Fragment Analyzer (Agilent, Santa Clara, USA). RNA-seq libraries were prepared using the TruSeq Stranded mRNA-seq reagents (Illumina) using 200ng of total RNA following the protocol provided by the supplier. The quality of the RNA-seq libraries was assessed on Fragment Analyzer (Agilent) and the libraries were quantified using the Qubit (Thermo Fisher). The libraries were sequenced on Illumina HiSeq 4000 using the 75-nucleotide paired end run configuration following the protocol provided by the supplier.

### Sample preparation for mass spectrometry proteotyping measurements

Samples were processed in blocks of 8 taking into consideration a balance between disease types and control samples. All other factors within a block were randomized. 230 samples were processed in three batches. For sample processing, aliquots of primary fibroblast (∼1e6 cells per vial, frozen in either DMEM or FBS or a mix of DMEM/FBS, plus 10% DMSO) were washed twice in ice-cold PBS (Gibco). After centrifugation for 5 minutes at 300g, the cells were resuspended in lysis buffer (Preomics) at a ratio of 1:1 (vol pellet/vol lysis buffer) and incubated at 95°C for 10 mins. Samples were sonicated in a vial tweeter (Hielscher Ultrasound Technology) at 4°C for 3 cycles with an amplitude 100%, power 80% during 30 secs. For each sample, the total protein concentration was estimated by nanodrop. 100 ug of protein lysate were further processed with the iST kit (Preomics). The purified peptides were resuspended in LCLoad buffer containing iRT peptides (Biognosys) at a concentration of 1 ug/ul.

### Spectral library generation

For spectral library generation, three times 24 samples (3×8 sample blocks) were pooled. Pooled sample batches were digested as described above. 100 μg of purified peptides were fractionated on a C18 column (YMC-Triart, C18, 3μm, 250 × 0.5 mm ID) according to pH on an Agilent HPLC 1260 system with a stepped 61 min gradient ranging from 95 % buffer A (20 mM ammonium formate acid/H2O) to 85% buffer B (20 mM ammonium formate/90 % ACN). 48 fractions were collected per sample and subsequently pooled to 24 fractions. Samples were resuspended in 5% ACN/0.1% FA and analyzed on a QExactive HF-X mass spectrometer (Thermo Fisher Scientific) in DDA mode. The same nLC 1200 configuration and mobile phase gradient elution conditions as for DIA were applied.

Full MS survey scans were acquired at a resolution of 60,000 with automatic gain control (AGC) target of 3e6 and a maximum injection time of 45 ms over a scan range of m/z 375-1500. A data-dependent top 12 method was used for HCD MS/MS with a normalized collision energy of 28 at a resolution of 15,000 and a fixed first mass of m/z 100. Precursor ions were isolated in a 1.4-Th window and accumulated to reach an AGC target value of 1e5 with a maximum injection time of 22 ms. Precursor ions with a charge state of 1 and 6 as well as isotopes were excluded for fragmentation. Dynamic exclusion was set to 15s.

DDA raw files were processed with Proteome Discoverer (v. 2.2) using a human UniProt database (release 201804) together with iRT peptides (Biognosys) and common contaminants. The processing workflow consisted of SequestHT ^42^ and Amanda ^43^ nodes coupled with Percolator ^44^. The following search parameters were used for protein identification: (i) a peptide mass tolerance of 10 ppm; (ii) an MS/MS mass tolerance of 0.02 Da; (iii) fully tryptic peptide search with up to two missed cleavages were allowed; (iv) carbamidomethylation of cysteine was set as fixed modification, methionine oxidation and protein N-term acetylation were set as variable modifications. Percolator was set at max delta Cn 0.05, with target FDR strict 0.01 and target FDR relaxed 0.05. The spectral library from Proteome Discoverer was imported into Spectronaut v12 (Biognosys, Schlieren, Switzerland) using standard parameters with 0.01 peptide spectrum match (PSM) FDR.

### DIA-MS setup and data analysis

For DIA analysis samples were measured on a Q Exactive HF mass spectrometer (Thermo Fisher Scientific). Mobile phase A consisted of HPLC-grade water with 0.1% (v/v) formic acid, and mobile phase B consisted of HPLC-grade ACN with 20% (v/v) HPLC-grade water and 0.1% (v/v) formic acid. Peptide separation was carried out on an ES806, 2 µm, 100 Å, 150 µm i.d. x 150 mm, C18 EASY-Spray column (Thermo Fisher Scientific) at a temperature of 50°C. For LC-MS/MS analyses, 2 *μ*g of each sample were loaded onto the column via an Easy-nLC 1200 system (Thermo Fisher Scientific). Samples were loaded at 4 µL/min with 100% mobile phase A for 5 min. Peptide elution was performed using the following gradient: i) 2% to 8% mobile phase B in 4 min, ii) 8% to 32% mobile phase B in 49 min, iii) 32% to 60 % mobile phase B in 1 min, and iv) ramp to 98% mobile phase B in 1 min at 2 µL/min.

For DIA-Acquisition on a Q Exactive HF mass spectrometer, we applied a DIA method published elsewhere ^45^. In short, we performed an MS1 scan over a mass range of m/z 400-1210 at a resolution of 120,000 with an AGC target value of 3e6 and with a maximum injection time of 50 ms. For MS/MS scans, resolution was at 0,000 with an AGC target value of 1e6 and with “Auto” maximum injection time. Precursor ions were isolated within a 15-Th window and fragmented by HCD with normalized collision energy 28. 54 MS/MS scan windows were defined, interspersed every 18 scans with an MS1 scan.

DIA data Analysis was performed in Spectronaut v12 (Biognosys) using standard parameters. For identification, a Qvalue cut-off of 0.01 was applied on the precursor as well as on the protein level. The MS1 area was selected for quantification. Quantification parameters were set to mean peptide quantity for major group quantity, the top 3 peptides were selected for protein quantity calculation. Data filtering was set to Qvalue sparse, with no imputation. Cross-run normalization was set to local. The protein report for downstream analysis contained information report about PG.ProteinAccessions, PG.ProteinDescriptions, PG.ProteinNames. PG.Qvalue and PG.Quantity.

### Selection of primary fibroblasts for polar metabolomics

To select cell lines for metabolomics, we opted for a balanced design with 10 MMUT deficient cell lines and 10 control lines. MMUT deficient lines were picked to show over-expression of GLUD1 and under-expression of OGDH whereas the control lines were chosen to show the reverse pattern. We fitted a mixed effects model with PI+ as response, two fixed effects for GLUD1 and OGDH expression, and a random effect with the same covariance structure as the proteomics data after column and row normalization. From the MMUT deficient and control cell lines, we chose 10 with the lowest predicted value of PI+ and 10 with the highest predicted value respectively. The top ten ranked MMUT deficient (MMA014, MMA92, MMA042, MMA67, MMA93, MMA104, MMA013, MMA030, MMA138, MMA036) and the last 10 ranked control primary fibroblasts (MMA219, MMA221, MMA227, MMA222, MMA213, MMA230, MMA226, MMA228, MMA225, MMA215) were selected and cultured as described above. Four primary fibroblasts in each group did not meet growth criteria, most likely due to the long freezing period, and were excluded (MMA014, MMA92, MMA042, MMA013, MMA219, MMA221, MMA227, MMA226). Six primary fibroblast lines per group were selected for the polar metabolomics experiment.

### Quality assessment of WGS, RNA-seq and DIA-MS data

Overall quality assurance tests revealed a mean of high-quality aligned genomic reads of 8.7×10^8^ at a median genomic coverage of >38-fold (**Extended data Fig. 1b**). A median of 3.74 million SNVs were called using the Genome Analysis Toolkit ^46^ and DeepVariant ^47^. RNA-seq data showed a median Phred score of >36.3 at three and more cycles (**Extended data Fig. 1c**), while proteomics data showed a high reproducibility with 2218 proteins detected in at least 75% of samples (**Extended data Fig. 1d**). For nine of the 230 samples RNA extraction yielded insufficient nucleic acid amounts to proceed with transcriptome sequencing; hence, these datasets were excluded from all further analysis (transcriptomics data of sample IDs 22, 54, 59, 78, 89, 109, 123, 207, 221).

### Fibroblast sample preparation for polar metabolomics

100,000 cells per well were seeded in a 6 well plate and grown for 48 hours. Media was removed and cells washed with 2 mL per well of 150 mM ammonium hydrogen carbonate (NH_4_HCO_3_) at pH 7.4 twice. The whole plate was flash-frozen in liquid nitrogen for 20 seconds and then stored at -80°C. Metabolites were extracted by putting the plate on dry ice and adding 400 ul of cold (−20°C) 40:40:20 acetonitrile:methanol:water and incubated at -20°C for 10 minutes. Supernatant was collected. Added another 400 ul of 40:40:20 acetonitrile:methanol:water and incubated at -20°C for 10 minutes. Plates were put on dry ice and cells were scraped mechanically. The supernatant including the scraped cells were collected in the same tube as during the first extraction step. Collection tubes were centrifuged at 13,000 rpm for 2 minutes at 4°C. Supernatants were stored at -20°C prior to metabolomics analysis.

### Polar metabolomics in patient-derived fibroblasts

Untargeted metabolite profiling was performed using flow injection analysis on an Agilent 6550 QTOF instrument (Agilent, Santa Clara, CA) using negative ionization, 4 GHz high resolution acquisition, and scanning in MS1 mode between m/z 50-1000 at 1.4 Hz ^48^. The solvent was 60:40 isopropanol:water supplemented with 1 mM NH4F at pH 9.0, as well as 10 nM hexakis(1H, 1H, 3H-tetrafluoropropoxy)phosphazine and 80 nM taurochloric acid for online mass calibration. The seven batches were analyzed sequentially. Within each batch, the injection sequence was randomized. Data was acquired in profile mode, centroided and analyzed with Matlab (The Mathworks, Natick). Missing values were filled by recursion in the raw data. Upon identification of consensus centroids across all samples, ions were putatively annotated by accurate mass and isotopic patterns. Starting from the HMDB v4.0 database, we generated a list of expected ions including deprotonated, fluorinated, and all major adducts found under these conditions. All formulas matching the measured mass within a mass tolerance of 0.001 Da were enumerated. As this method does not employ chromatographic separation or in-depth MS2 characterization, it is not possible to distinguish between compounds with identical molecular formula. The confidence of annotation reflects Level 4 but – in practice – in the case of intermediates of primary metabolism it is higher because they are the most abundant metabolites in cells. The resulting data matrix included 1809 ions that could be matched to deprotonated metabolites listed in HMDB. All m/z peaks that remained unmatched or were associated to adducts or heavy isotopomers were discarded.

### Mouse care and handling

Ethics approval was obtained prior to the study from the Cantonal Veterinary Office Zurich under the license number 202/2014. Mice were kept under standard conditions, i.e. single-ventilated cages with controlled humidity and room temperature of 22°C. Generation of the *Mmut*-p.Met698Lys variant model and crossing with a *Mmut*-ko/wt model was done as previously described ^23^.

### Harvesting of mouse tissues

Urine was collected in the morning after one night in a metabolic cage. The sediment was removed, and supernatant was flash frozen in liquid nitrogen. Tissue samples were harvested from mice aged 58 to 63 days. Animals were anesthetized by sevoflurane. Portal blood was taken and kept on ice to coagulate, centrifuged at 4°C and snap frozen in liquid nitrogen directly after. Liver, kidneys, heart, and brain were harvested and snap-frozen in liquid nitrogen. After the procedure, the mice were directly euthanized by cervical dislocation. All samples were stored at -80°C prior to analysis.

### Metabolomics in mouse tissues

The mouse body fluid and tissue samples derived from five *Mmut*-ki/wt and five *Mmut*-ko/ki mice were harvested as described above and prepared as previously published ^49^. Sample analysis using liquid chromatography-mass spectrometry was performed as previously published ^50^. Ions were annotated to metabolites based on exact mass to the KEGG database ^51^ considering [M-H+] and 0.01 Da mass accuracy. Significantly changing ions between mutant and control conditions were identified using a two-sample t-test. Pathway analysis was performed using annotated ion list ranked by p-value significance. Pathway enrichments were calculated using KEGG metabolic pathway definitions and a hypergeometric test.

### Transcriptomics in mouse brains

Brain tissue samples were harvested as described above. Four mice per genotype groups *Mmut*-ki/wt and *Mmut*-ko/ki were used. RNA was purified using DNAse kit (Qiagen, No. 79254) together with QIAmp RNA Blood Mini Kit (Qiagen, No. 52304). RNA-seq reads were aligned with the STAR-aligner ^52^. As reference we used the Ensembl mouse genome build GRCm38. Gene expression values were computed with the function featureCounts from the R package Rsubread ^53^.

### CRISPR gene-editing experiments

CRISPR-Cas9 editing was performed as described ^54^. Cas9 protein was provided as a plasmid (PX459-V2.0, Addgene, 62988) and guide RNA as gBLOCKS ^55^ (IDT Technologies). 293T cells were transfected using the Neon transfection system (Thermo Fisher Scientific) containing 100,000 cells, 0.6 µg of Cas9 plasmid and 600 ng of guide RNA following manufacturer’s instructions. 48 h after transfection, cells were collected, diluted to 1 cell/100 µl and transferred to a 96-well plate at 100 µl/well for clonal selection. Correct clones were confirmed by Sanger sequencing of genomic DNA.

### Western blotting

Lysates of 293T cell lines were mixed with RIPA lysis buffer and 4X Laemmli buffer to obtain a concentration of 1 μg/μl protein. Samples were incubated at 96°C for 5 minutes. 20 μl of each sample was loaded onto a 10% SDS page gel. Proteins were separated by electrophoresis, transferred with a semi-dry method onto a nitrocellulose membrane (Whatman, GE Healthcare), blocked at room temperature for 2 hours with buffer A (5% skimmed milk, 1.2% w/v Tris-base, 9% w/v NaCl, 0.2% Tween 20, pH 7.6), incubated with primary antibodies dissolved in buffer A overnight at 4°C, and detected using secondary antibodies in buffer A. Primary antibodies used were probing for the following proteins: MMUT (Abcam, ab67869, 1:1000, host: mouse), OGDH (Atlas antibodies, HPA020347, 1:500, host: rabbit), GLUD (Abcam, ab166618, 1:2000, host: rabbit), Beta-actin (Sigma, A1978, 1:5000, host: mouse). Secondary antibodies used were Anti-rabbit HRP (Santa Cruz, sc-2357, 1:5000, host: mouse), Anti-mouse HRP (Santa Cruz, sc-516102, 1:5000, host: goat).

### MMUT enzyme activity assay

Confluent cells from a T75 were detached using trypsin, the cell suspension centrifuged at 250 *g*, the pellet washed two-times 1 mL in PBS and the supernatant discarded. Homogenates were obtained by resuspension of cell pellets in 150 μL of 5 mM KH_2_PO_4_ (pH 7.4) followed by sonication on ice with 2 pulses for 15 seconds using the microprobe of an XL-2000 (Microson, Qsonica, Newtown, CT, USA) with amplitude set to 2 microns. The homogenate was then diluted 1:4 in a final concentration of 50 μM adenosylcobalamin (Sigma C0884), 1 mM methylmalonyl-CoA (Sigma M1762) and 0.1 M assay buffer KH_2_PO_4_. The enzymatic reaction was performed at 37°C for 30 min. Subsequently, the reaction was stopped, and succinyl-CoA hydrolysed by addition of 500 mM KOH and incubation at 37°C for 15 min. The assay mixture was neutralized with HClO_4_ at a final concentration of 400 mM, the sample centrifuged at 16,000 *g* and the supernatant discarded. Precipitates were frozen at -20 degrees until further analysis. For mass spectrometry, the MMUT-KO sample was diluted 1:2, the WT sample was diluted 1:50 in a dilution solution (Recipe, MS5021) and then precipitated with 4 parts precipitation reagent with internal standard (Recipe, MS5112). After centrifugation (5min, 16’000 *g*) the supernatant was transferred into a fresh vial. Final succinate determination was performed by HPLC separation and electrospray tandem mass spectrometry (ESI-MS/MS) detection (SCIEX TripleQuad 5500 LC-MS/MS System).

### KGDH enzyme activity assays

Assay of oxoglutarate dehydrogenase enzyme activity was performed in 293T cell clones according to the manufacturer’s instructions (Sigma-Aldrich, St. Louis, USA, catalogue number MAK189) using the plate reader Victor Nivo by PerkinElmer.

### Glutamine tracing studies

293T cells were cultured on coverslips in DMEM (Gibco, catalog number: 11960044) without L-glutamine, sodium pyruvate and HEPES supplemented with 10% FBS, 1% antibiotic-antimycotic (Gibco), and [U-^13^C]glutamine (final concentration 4 mM) (Sigma-Aldrich, catalog number: 605166) for 4 hours. Medium was then removed, coverslips were quickly dipped into sterile double-distilled water at 37°C and quenched in 80% methanol at -80°C. Cells were scrapped in methanol, collected, and centrifuged at 15’000 g for 15 minutes at 4°C. Supernatants were collected, snap frozen in liquid nitrogen, and stored at -80°C prior to LC/MS analysis.

300 µl of sample was lyophilized overnight until dry and resolubilized in 200 µl loading buffer (water, 0.5% formic acid) in narrow-bottom 96-deep well plates on shaker (800 rpm, 15°C, 10 min) for LC-MS injection. Metabolites were separated using a ACQUITY UPLC HSS T3 1.8 µm, 100 × 2.1 mm I.D. column (Waters, Massachusetts, US) and eluted using the following gradient from solvent A (water, 5 mM ammonium formate, 0.1% formic acid) to solvent B (methanol, 5 mM ammonium formate, 0.1% formic acid) as follows: 2 minutes at 0% B, 2-3.5 mins to 4% B, 3.5-10 mins to 45% B, 10-12 mins to 70% B, 12-13.5s min to 100% B, with a isocratic plateau at 100% B for 2 mins to 15.5 mins, and from 15.5-16.5 mins to 0%B. After each run the column was re-equilibrated for 8 mins at 100% A with a constant flow rate of 0.4ml/min.

Mass spectra were acquired using a heated electro-spray ionization (HESI) source of a Q-Exactive high resolution, accurate mass spectrometer (Thermo Scientific, Waltham, MS, USA). Mass spectra were recorded in positive and negative mode with the MS detector in full-scan mode (Full-MS) in the scan-range 50 to 750 m/z with an automatic gain control target of 1e6, an Orbitrap resolution of 70,000, and a maximum injection time of 80 ms. Peaks were integrated with Xcalibur (version 4.0.27.19, Thermo Fisher Scientific) using windows of 0.01 m/z and 20 seconds for retention time as previously determined using a library of standards.

HESI parameters: sheath gas flow rate 35 arbitrary units (AU), auxiliary gas flow rate 35 AU, sweep gas flow rate 2 AU, spray voltage 3.5 kV, capillary temperature 350°C, aux gas heater temperature 350°C. Detector settings for full MS: In-source CID 0.0 eV, µscans = 1, resolution = 70,000, AGC target 1e6, max IT = 35 ms, spectrum data type, profile. Integration parameters: ICIS Peak Integration, nearest RT, smoothing points 3, baseline window 40, area noise factor 3, peak noise factor 70, minimum peak height 3.0.

Data preprocessing included missing value imputation and normalization to internal standards [^2^H]_3_-creatine and [^2^H]_4_-citric acid for positive and negative mode respectively.

### Affinity capture mass spectrometry

293T cells (ATCC #CRL-3216; Manassas, WV) were grown in Dulbecco’s Modified Eagle Medium (Gibco, Carlsbad, CA) supplemented with 10% fetal bovine serum (Gibco) and antibiotics (GE Healthcare). Transient transfection of pCDNA3-C-Flag-LIC constructs was performed using Lipofectamine 3000 (Thermo Fisher Scientific) according to manufacturer’s instructions. 48 h after transfection, cells were crosslinked using 0.5% paraformaldehyde (PFA, Sigma-Aldrich) in PBS (Gibco) for 10 min at RT, the reaction was quenched with 1.25 M glycine/PBS (Sigma-Aldrich) for 10 min at 4°C, cells were centrifuged for 5 min at 2,000×*g* at 4°C, and the pellet resuspended in lysis buffer (1% Nonidet P-40, 0.5% deoxycholine, 150 mM NaCl, 50 mM Tris–HCl, pH7.5, all Sigma-Aldrich). Pre-cleared cell extracts were immunoprecipitated with anti-flag M2 (F3165, Sigma– Aldrich) using Dynabeads Protein G (Thermo Fisher Scientific) according to the manufacturer’s instructions. Following washing, peptides were released by trypsin (100 ng/µl in 10 mM HCl) and supernatants collected, dried, dissolved in 0.1% formic acid.

All affinity-captured samples were measured on a Q Exactive mass spectrometer (Thermo Fisher Scientific) with a MS1 resolution of 70,000 and an AGC target of 3e6 and a maximum injection time of 100 ms over a scan range of m/z 350-1500. A data-dependent top 12 method was used for HCD MS/MS with a normalized collision energy of 25 at a resolution of 35,000. Precursor ions were isolated in a 1.2-Th window with an AGC target value of 1e5 with a maximum injection time of 120 ms. Dynamic exclusion was set to 40s.

Samples were analyzed using Mascot (Matrix Science, London, UK; version 2.6.2) with the SwissProt database (download date 20190204) assuming trypsin with at maximum two miscleavages. Mascot was searched with a fragment ion mass tolerance of 0.030 Da and a parent ion tolerance of 10.0 PPM. Oxidation of methionine was specified as a variable modification. Scaffold (version Scaffold_5.1.2, Proteome Software Inc., Portland, OR) was used to validate MS/MS based peptide and protein identifications. Peptide identifications were accepted if they could be established at greater than 95.0% probability by the Scaffold Local FDR algorithm. Protein identifications were accepted if they could be established at greater than 99.0% probability and contained at least 2 identified peptides.

### Data analysis

Data analysis was performed using R version 4.1.0. For the global data layer inspection we used the MOFA package version 1.3.1 ^22^, the MASS package version 7.3-54 ^56^, the fgsea package version 1.18.0 ^57^. Gene enrichment analysis was performed using gene sets downloaded from http://www.gsea-msigdb.org/gsea/msigdb/index.jsp “MSigDB Collections” on 28 December 2020. Circos including chord plots were created using the circlize package version 0.4.13 ^58^. The Uniprot portal was accessed on 24 February 2021 at 4 PM to scrape protein localization data. The code for all the analyses and generation of figures is hosted on a repository on the GitHub platform under https://github.com/pforny/MMAomics.

### Genetic variant investigation approach

Short variant calling was carried out with GATK and Deep Variant algorithms, and annotated with annovar ^59^. CNVs were called with CNVnator ^60^ with a bin size of 100 and standard parameters, and annotated with AnnotSV ^61^. Variation in the *MMUT* gene was investigated first. When no genetic cause for the phenotype was identified with this approach (two inactivating/damaging events in *MMUT*), other genes known to be involved in MMA (based on literature reports) were investigated as a virtual gene panel. When no genetic cause was found in the two previous steps, genes highlighted by mutational burden (genes harboring pathogenic variants across the cohort in an autosomal recessive pattern in two or more individuals) were investigated. Finally, all samples and controls were used to run OUTRIDER ^19^, and genes highlighted as expression outliers associated with phenotypes overlapping MMA were analyzed to either confirm the identified damaging variants, or to further explore damaging variation in them.

Variants were prioritized with the following approach: First, any coding variant (excluding synonymous variants) with a GnomAD frequency across all represented populations <0.01, in homozygosity or compound heterozygosity with another relevant variant, and supported by at least two forward and two reverse reads and at least 8 reads coverage, were evaluated. Second, all variants categorized by the automatic application of the ACMG criteria ^62^ by InterVar ^63^, or classified in ClinVar ^64^ as “pathogenic” or “likely pathogenic”, in homozygosity or compound heterozygosity with another relevant variant, were considered and evaluated. Third, variants with a dbscSNV_ADA or dbscSNV_RF scores >0.6 in the annovar annotation using the database prepared and described previously ^65^ were evaluated.

For CNVs, individuals with a single heterozygous variant or no variation in *MMUT* and the other genes of interest were investigated for the presence of relevant CNVs that could explain their phenotype ^59^.

## References

1. Argmann, C. A., Houten, S. M., Zhu, J. & Schadt, E. E. A Next Generation Multiscale View of Inborn Errors of Metabolism. Cell Metab 23, 13–26 (2016).

2. Garrod, ArchibaldE. The Croonian Lectures ON INBORN ERRORS OF METABOLISM. The Lancet 172, 1–7 (1908).

3. Ferreira, C. R., Rahman, S., Keller, M. & Zschocke, J. An international classification of inherited metabolic disorders (ICIMD). Journal of Inherited Metabolic Disease 44, 164–177 (2021).

4. Rahman, S. Mitochondrial disease in children. J Intern Med 287, 609–633 (2020).

5. Hirano, M., Emmanuele, V. & Quinzii, C. M. Emerging therapies for mitochondrial diseases. Essays Biochem 62, 467–481 (2018).

6. 100,000 Genomes Project Pilot Investigators et al. 100,000 Genomes Pilot on Rare-Disease Diagnosis in Health Care - Preliminary Report. N Engl J Med 385, 1868–1880 (2021).

7. Palmer, E. E. et al. Diagnostic Yield of Whole Genome Sequencing After Nondiagnostic Exome Sequencing or Gene Panel in Developmental and Epileptic Encephalopathies. Neurology 96, e1770–e1782 (2021).

8. Schon, K. R. et al. Use of whole genome sequencing to determine genetic basis of suspected mitochondrial disorders: cohort study. BMJ e066288 (2021) doi:10.1136/bmj-2021-066288.

9. Cummings, B. B. et al. Improving genetic diagnosis in Mendelian disease with transcriptome sequencing. Sci Transl Med 9, eaal5209 (2017).

10. Kremer, L. S. et al. Genetic diagnosis of Mendelian disorders via RNA sequencing. Nat Commun 8, 15824 (2017).

11. Gonorazky, H. D. et al. Expanding the Boundaries of RNA Sequencing as a Diagnostic Tool for Rare Mendelian Disease. Am J Hum Genet 104, 466–483 (2019).

12. Frésard, L. et al. Identification of rare-disease genes using blood transcriptome sequencing and large control cohorts. Nat Med 25, 911–919 (2019).

13. Froese, D. S. & Gravel, R. A. Genetic disorders of vitamin B12 metabolism: eight complementation groups--eight genes. Expert Rev Mol Med 12, e37 (2010).

14. Forny, P., Froese, D. S., Suormala, T., Yue, W. W. & Baumgartner, M. R. Functional characterization and categorization of missense mutations that cause methylmalonyl-CoA mutase (MUT) deficiency. Hum Mutat 35, 1449–1458 (2014).

15. Froese, D. S. et al. Structures of the Human GTPase MMAA and Vitamin B12-dependent Methylmalonyl-CoA Mutase and Insight into Their Complex Formation*,. Journal of Biological Chemistry 285, 38204–38213 (2010).

16. Ruetz, M. et al. Itaconyl-CoA forms a stable biradical in methylmalonyl-CoA mutase and derails its activity and repair. Science 366, 589–593 (2019).

17. Jost, M., Cracan, V., Hubbard, P. A., Banerjee, R. & Drennan, C. L. Visualization of a radical B12 enzyme with its G-protein chaperone. Proc Natl Acad Sci U S A 112, 2419–2424 (2015).

18. Froese, S. & Baumgartner, M. R. Lysosomal Vitamin B12 Trafficking. in Ion and Molecule Transport in Lysosomes (CRC Press, 2020).

19. Brechtmann, F. et al. OUTRIDER: A Statistical Method for Detecting Aberrantly Expressed Genes in RNA Sequencing Data. The American Journal of Human Genetics 103, 907–917 (2018).

20. Forny, P. et al. Guidelines for the diagnosis and management of methylmalonic acidaemia and propionic acidaemia: First revision. Journal of Inherited Metabolic Disease 44, 566–592 (2021).

21. Hörster, F. et al. Prediction of outcome in isolated methylmalonic acidurias: combined use of clinical and biochemical parameters. Journal of Inherited Metabolic Disease 32, 630 (2009).

22. Argelaguet, R. et al. Multi-Omics Factor Analysis-a framework for unsupervised integration of multi-omics data sets. Mol Syst Biol 14, e8124 (2018).

23. Forny, P. et al. Novel Mouse Models of Methylmalonic Aciduria Recapitulate Phenotypic Traits with a Genetic Dosage Effect. J Biol Chem 291, 20563–20573 (2016).

24. Lamparter, D., Marbach, D., Rueedi, R., Bergmann, S. & Kutalik, Z. Genome-Wide Association between Transcription Factor Expression and Chromatin Accessibility Reveals Regulators of Chromatin Accessibility. PLOS Computational Biology 13, e1005311 (2017).

25. Rath, S. et al. MitoCarta3.0: an updated mitochondrial proteome now with sub-organelle localization and pathway annotations. Nucleic Acids Res 49, D1541–D1547 (2021).

26. Nusinow, D. P. et al. Quantitative Proteomics of the Cancer Cell Line Encyclopedia. Cell 180, 387–402.e16 (2020).

27. Williams, E. G. et al. Systems proteomics of liver mitochondria function. Science 352, aad0189– aad0189 (2016).

28. Heusel, M. et al. Complex-centric proteome profiling by SEC-SWATH-MS. Mol Syst Biol 15, e8438 (2019).

29. Buescher, J. M. et al. A roadmap for interpreting (13)C metabolite labeling patterns from cells. Curr Opin Biotechnol 34, 189–201 (2015).

30. Chen, Q. et al. Rewiring of Glutamine Metabolism Is a Bioenergetic Adaptation of Human Cells with Mitochondrial DNA Mutations. Cell Metabolism 27, 1007–1025.e5 (2018).

31. Zhang, J. et al. 13C isotope-assisted methods for quantifying glutamine metabolism in cancer cells. Methods Enzymol 542, 369–389 (2014).

32. Liu, A. et al. From expression footprints to causal pathways: contextualizing large signaling networks with CARNIVAL. npj Syst Biol Appl 5, 1–10 (2019).

33. Lewis, J. E., Forshaw, T. E., Boothman, D. A., Furdui, C. M. & Kemp, M. L. Personalized Genome-Scale Metabolic Models Identify Targets of Redox Metabolism in Radiation-Resistant Tumors. Cell Systems 12, 68–81.e11 (2021).

34. Wang, H. et al. Genome-scale metabolic network reconstruction of model animals as a platform for translational research. PNAS 118, (2021).

35. Sloan, J. L. et al. Exome sequencing identifies ACSF3 as a cause of combined malonic and methylmalonic aciduria. Nat Genet 43, 883–886 (2011).

36. Karczewski, K. J. & Snyder, M. P. Integrative omics for health and disease. Nat Rev Genet 19, 299–310 (2018).

37. Vockley, J. et al. Effects of triheptanoin (UX007) in patients with long-chain fatty acid oxidation disorders: Results from an open-label, long-term extension study. J Inherit Metab Dis 44, 253– 263 (2021).

38. Longo, N. et al. Anaplerotic therapy in propionic acidemia. Mol Genet Metab 122, 51–59 (2017).

39. Willard, H. F., Ambani, L. M., Hart, A. C., Mahoney, M. J. & Rosenberg, L. E. Rapid prenatal and postnatal detection of inborn errors of propionate, methylmalonate, and cobalamin metabolism: a sensitive assay using cultured cells. Hum Genet 34, 277–283 (1976).

40. Baumgartner, R. Activity of the cobalamin-dependent methylmalonyl-CoA mutase. The Cobalamins: Methods in Hematology. Churchill Livingstone, Edinburgh, New York 181–195 (1983).

41. Causey, A. G. & Bartlett, K. A radio-HPLC assay for the measurement of methylmalonyl-CoA mutase. Clinica Chimica Acta 139, 179–186 (1984).

42. Eng, J. K., McCormack, A. L. & Yates, J. R. An approach to correlate tandem mass spectral data of peptides with amino acid sequences in a protein database. J Am Soc Mass Spectrom 5, 976–989 (1994).

43. Dorfer, V. et al. MS Amanda, a universal identification algorithm optimized for high accuracy tandem mass spectra. J Proteome Res 13, 3679–3684 (2014).

44. Brosch, M., Yu, L., Hubbard, T. & Choudhary, J. Accurate and sensitive peptide identification with Mascot Percolator. J Proteome Res 8, 3176–3181 (2009).

45. Xuan, Y. et al. Standardization and harmonization of distributed multi-center proteotype analysis supporting precision medicine studies. Nat Commun 11, 5248 (2020).

46. McKenna, A. et al. The Genome Analysis Toolkit: a MapReduce framework for analyzing next-generation DNA sequencing data. Genome Res 20, 1297–1303 (2010).

47. Poplin, R. et al. A universal SNP and small-indel variant caller using deep neural networks. Nat Biotechnol 36, 983–987 (2018).

48. Fuhrer, T., Heer, D., Begemann, B. & Zamboni, N. High-throughput, accurate mass metabolome profiling of cellular extracts by flow injection-time-of-flight mass spectrometry. Anal Chem 83, 7074–7080 (2011).

49. Want, E. J. et al. Global metabolic profiling of animal and human tissues via UPLC-MS. Nat Protoc 8, 17–32 (2013).

50. Abela, L. et al. Plasma metabolomics reveals a diagnostic metabolic fingerprint for mitochondrial aconitase (ACO2) deficiency. PLoS One 12, e0176363 (2017).

51. Kanehisa, M., Furumichi, M., Tanabe, M., Sato, Y. & Morishima, K. KEGG: new perspectives on genomes, pathways, diseases and drugs. Nucleic Acids Res 45, D353–D361 (2017).

52. Dobin, A. et al. STAR: ultrafast universal RNA-seq aligner. Bioinformatics 29, 15–21 (2013).

53. Liao, Y., Smyth, G. K. & Shi, W. The Subread aligner: fast, accurate and scalable read mapping by seed-and-vote. Nucleic Acids Res 41, e108 (2013).

54. Ran, F. A. et al. Genome engineering using the CRISPR-Cas9 system. Nat Protoc 8, 2281–2308 (2013).

55. Arbab, M., Srinivasan, S., Hashimoto, T., Geijsen, N. & Sherwood, R. I. Cloning-free CRISPR. Stem Cell Reports 5, 908–917 (2015).

56. Venables, W. N. & Ripley, B. D. Modern applied statistics with S. (Springer, 2002).

57. Korotkevich, G. et al. Fast gene set enrichment analysis. 060012 https://www.biorxiv.org/content/10.1101/060012v3 (2021) doi:10.1101/060012.

58. Gu, Z., Gu, L., Eils, R., Schlesner, M. & Brors, B. circlize Implements and enhances circular visualization in R. Bioinformatics 30, 2811–2812 (2014).

59. Wang, K., Li, M. & Hakonarson, H. ANNOVAR: functional annotation of genetic variants from high-throughput sequencing data. Nucleic Acids Res 38, e164 (2010).

60. Abyzov, A., Urban, A. E., Snyder, M. & Gerstein, M. CNVnator: an approach to discover, genotype, and characterize typical and atypical CNVs from family and population genome sequencing. Genome Res 21, 974–984 (2011).

61. Geoffroy, V. et al. AnnotSV: an integrated tool for structural variations annotation. Bioinformatics 34, 3572–3574 (2018).

62. Richards, S. et al. Standards and guidelines for the interpretation of sequence variants: a joint consensus recommendation of the American College of Medical Genetics and Genomics and the Association for Molecular Pathology. Genet Med 17, 405–424 (2015).

63. Li, Q. & Wang, K. InterVar: Clinical Interpretation of Genetic Variants by the 2015 ACMG-AMP Guidelines. Am J Hum Genet 100, 267–280 (2017).

64. Landrum, M. J. et al. ClinVar: improving access to variant interpretations and supporting evidence. Nucleic Acids Res 46, D1062–D1067 (2018).

65. Jian, X., Boerwinkle, E. & Liu, X. In silico prediction of splice-altering single nucleotide variants in the human genome. Nucleic Acids Res 42, 13534–13544 (2014).

