## Extended data for "Integrated multi-omics reveals anaplerotic insufficiency in methylmalonyl-CoA mutase deficiency"

#### Extended data Fig. 1 Historic context of sample collection and quality control measurements of multi-omics data.

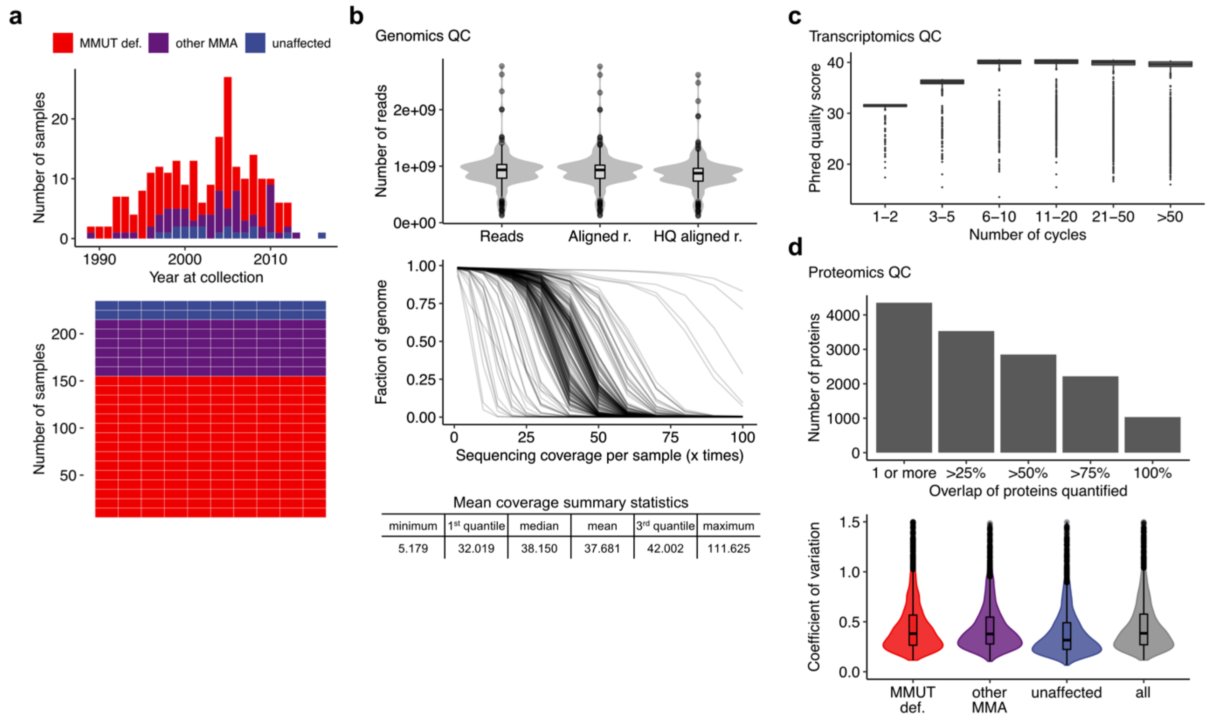

**a**, Histogram of fibroblast samples binned into their year at collection and waffle chart illustrating the different sample groups indicated by the color code. **b**, Violin plots illustrating average number of read per sample. HQ, high quality. Line plot (one line per sample) indicating genome coverage as quantitatively summarized in the below table. **c**, Boxplots indicating Phred quality scores at different number of cycles. **d**, Proteomics quality control illustrated by the percentage of overlapping proteins in variation coefficients in the respective sample groups.

### Extended data Fig. 2 Biochemical assessment of MMUT activity and propionate incorporation activity supports diagnosis of affected individuals.

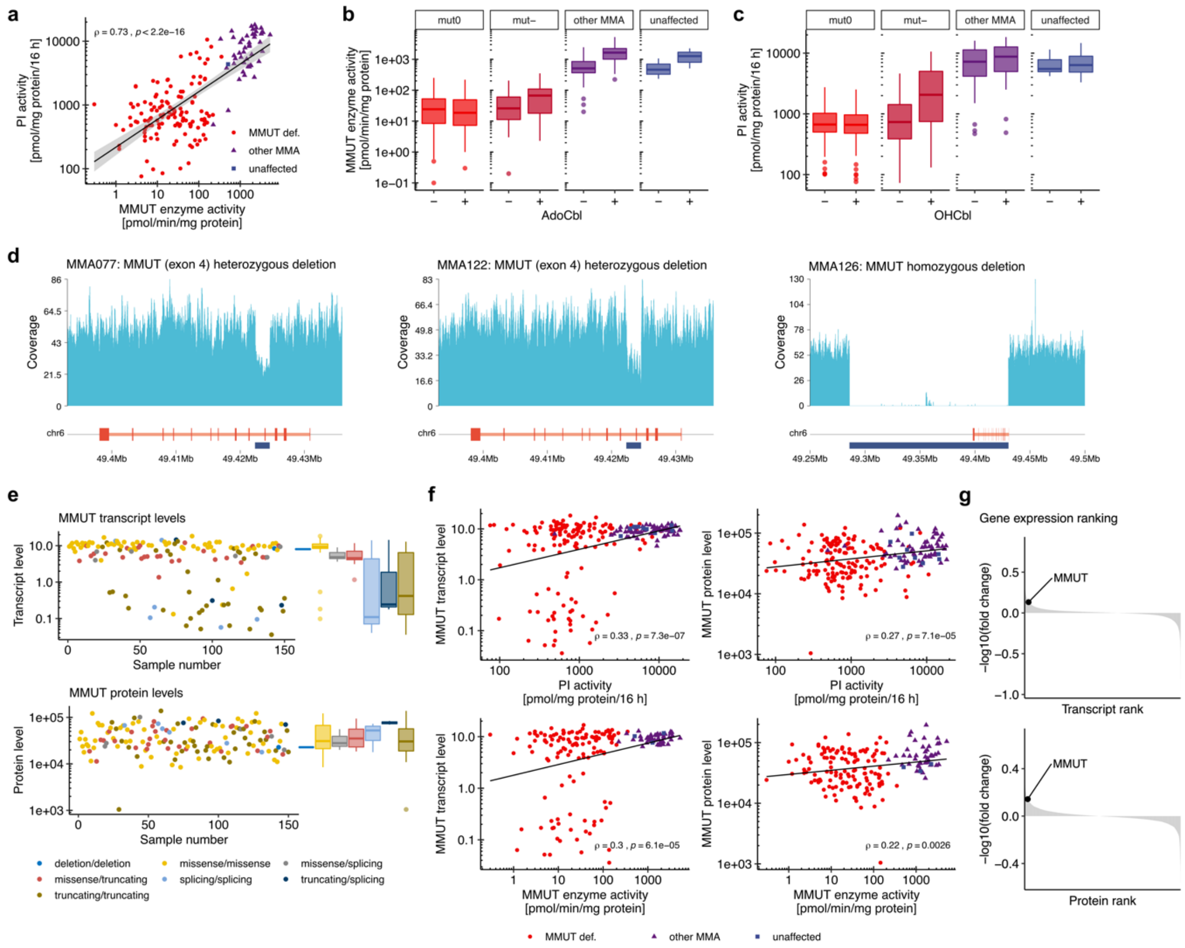

**a**, Scatter plot of maximal, i.e. supplemented with adenosylcobalamin (AdoCbl) or hydroxocobalamin (OHCbl), activity of the MMUT enzyme and the propionate incorporation assay. **b**, Boxplots of MMUT enzyme activity with and without AdoCbl supplementation. **c**, Boxplots of propionate incorporation activity with and without OHCbl supplementation. **d**, Copy number variants illustrated by read counts of specific locations of the *MMUT* gene for three specific samples. **e**, Scatter plots of MMUT transcript and protein levels of the MMUT-deficient samples. Each dot indicates one sample. Samples are grouped according to underlying biallelic genetic variation type of the *MMUT* gene. **f**, Regression plots of MMUT transcript and protein levels versus MMUT enzyme and propionate incorporation activity. **g**, Fold change of all transcripts and proteins respectively, when comparing the MMUT-deficient group versus the rest of the samples. Gene names are ranked according to the negative base 10 logarithm of the fold change. All linear regressions are calculated according to the Pearson method.

### Extended data Fig. 3 Expression outlier analysis reveals causative genes in specific disease samples.

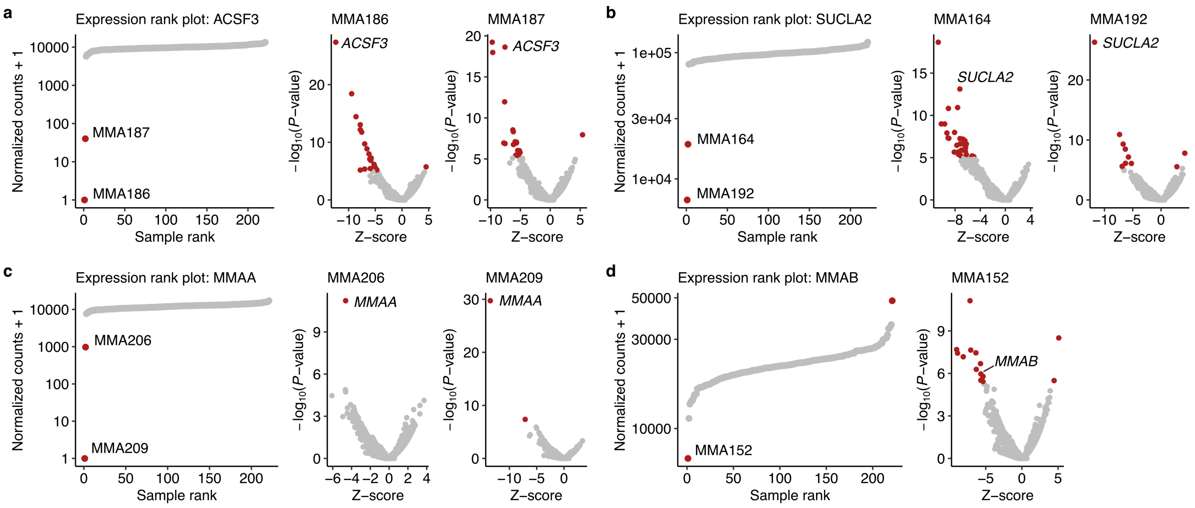

Expression rank plots for **a**, *ACSF3*, **b**, *SUCLA2*, **c**, *MMAA*, and **d**, *MMAB* and Z-score volcano plots for specific samples, applying the OUTRIDER R package.

### Extended data Fig. 4 The clinical severity score and propionate incorporation activity are associated with several phenotypic traits.

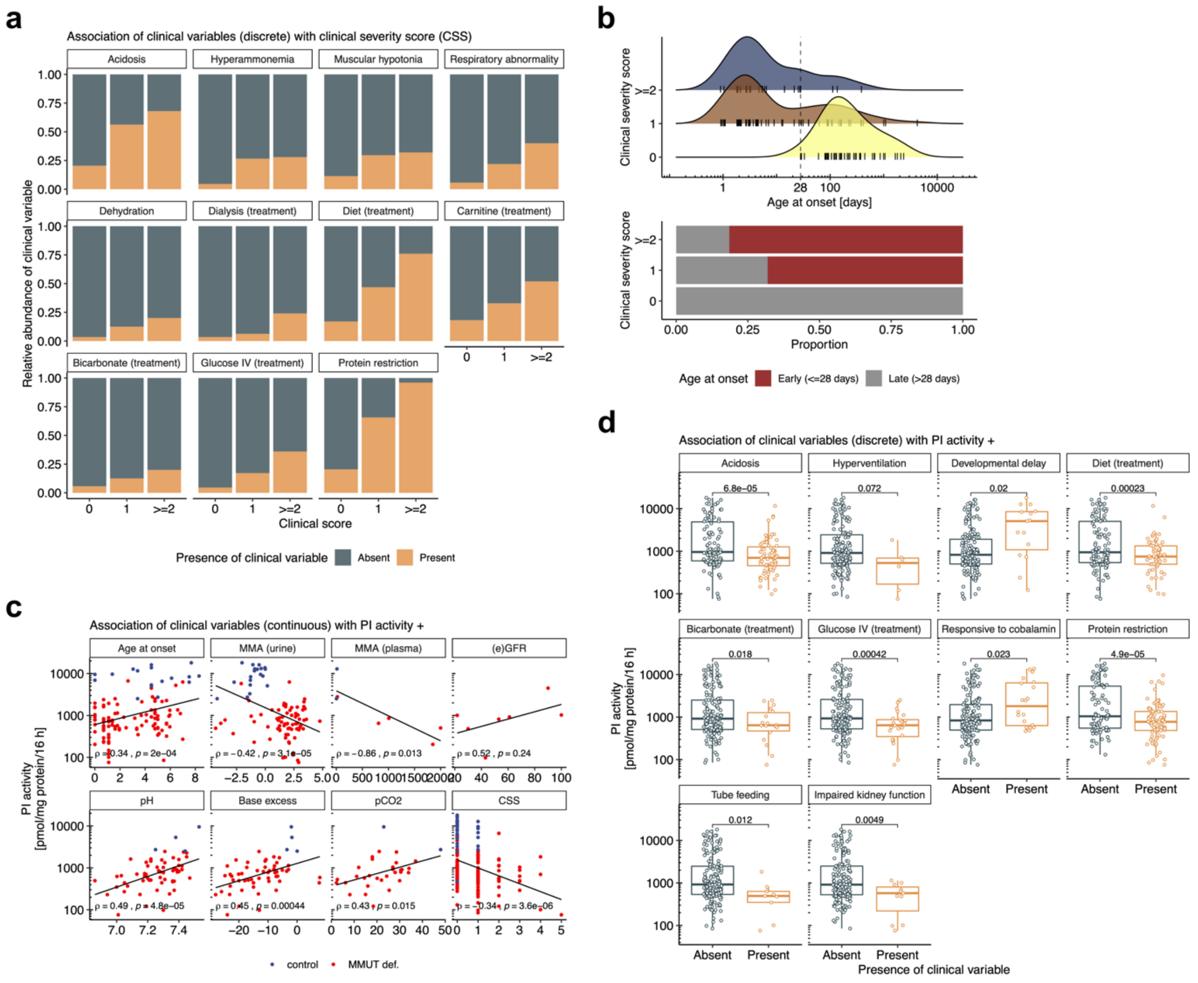

**a**, Proportional bar plots of the presence of absence of clinical parameters in relation to the clinical severity score. **b**, Age at onset in relation to the clinical severity score. Linear regression of various relationships of propionate incorporation activity to continuous (**c**) and discrete (**d**) phenotypic variables.

### Extended data Fig. 5 Global computational approaches to transcriptomics and proteotyping datasets were unable to stratify samples into disease and non-disease groups.

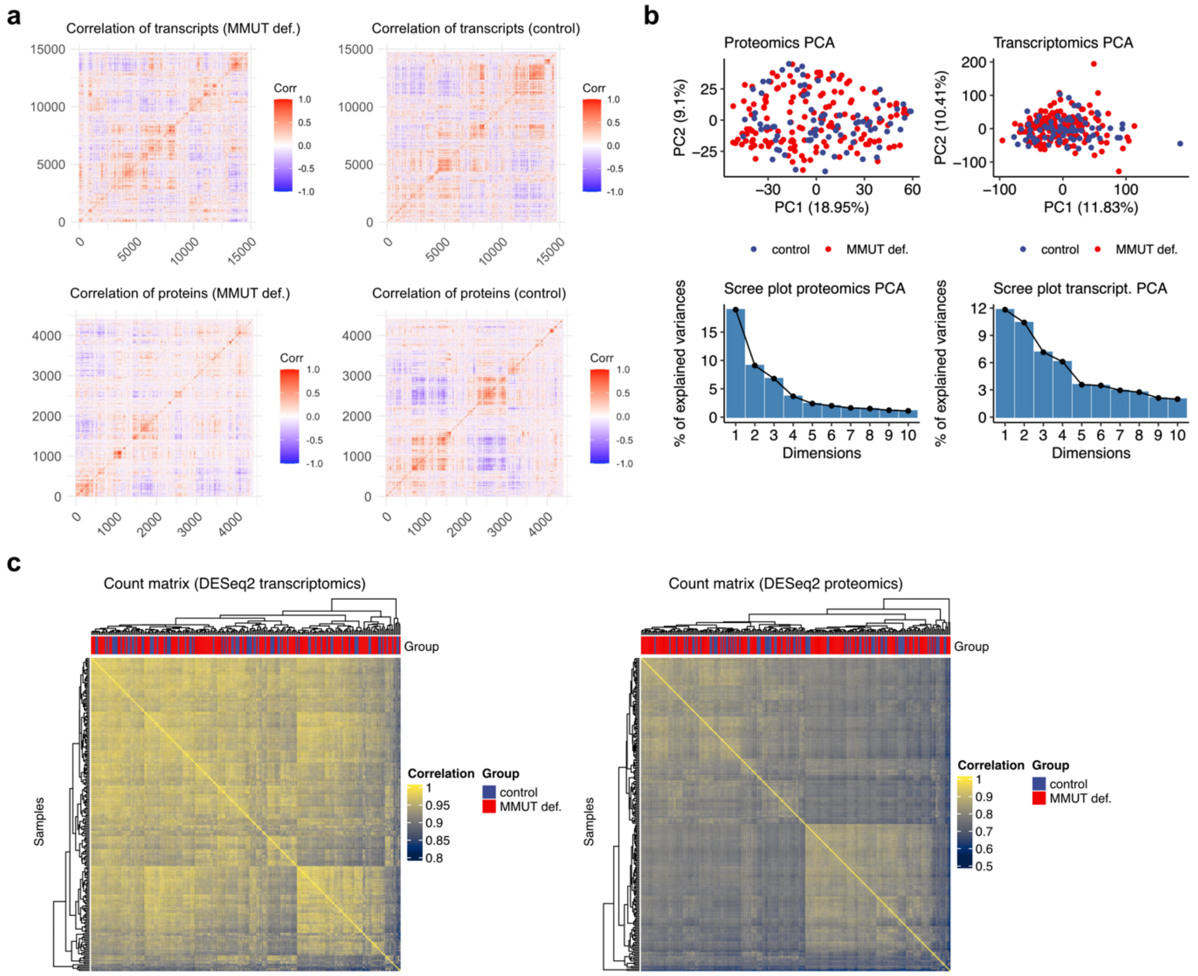

**a**, Pearson correlation matrices of all transcripts and proteins grouped by MMUT-deficient and control groups. **b**, Principal component analysis of transcriptomics and proteomics datasets with prior gene- and sample-wise scaling. **c**, Quality control heatmap of the differential expression analysis using the DESeq2 package.

#### Extended data Fig. 6 Transcriptomics analysis of mouse brain revealed sample clustering according to genotype.

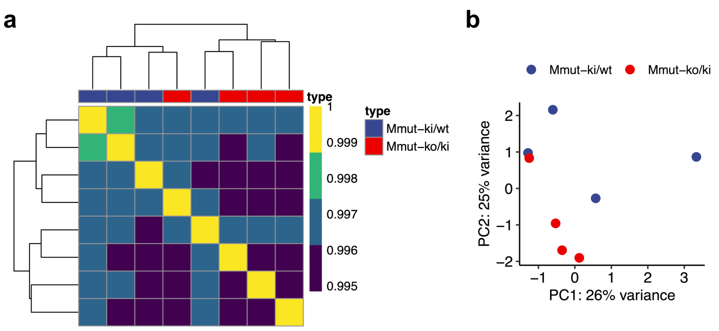

**a**, Quality control heatmap and, **b**, PCA plot based on differential expression analysis performed by applying the DESeq2 R package.

#### Extended data Fig. 7 Significantly dysregulated proteins were enriched for for mitochondrial localization.

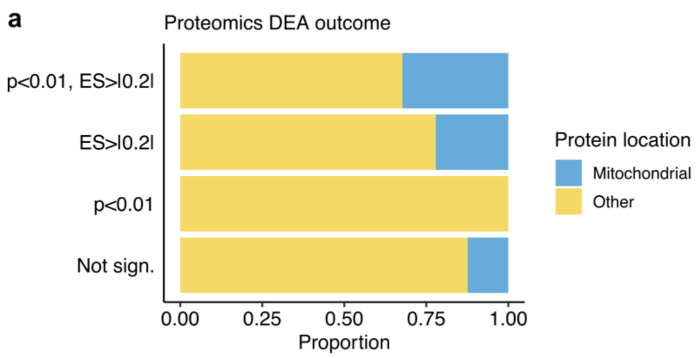

**a**, Proportions of proteins localized to the mitochondria (according to MitoCarta 3.0) split according to the result of the differential expression analysis (Fig. 4b); ES, effect size.

### Extended data Fig. 8 Transcript-protein and protein-protein correlation analysis illustrates coordinated regulation of MMUT with most TCA transcripts and proteins.

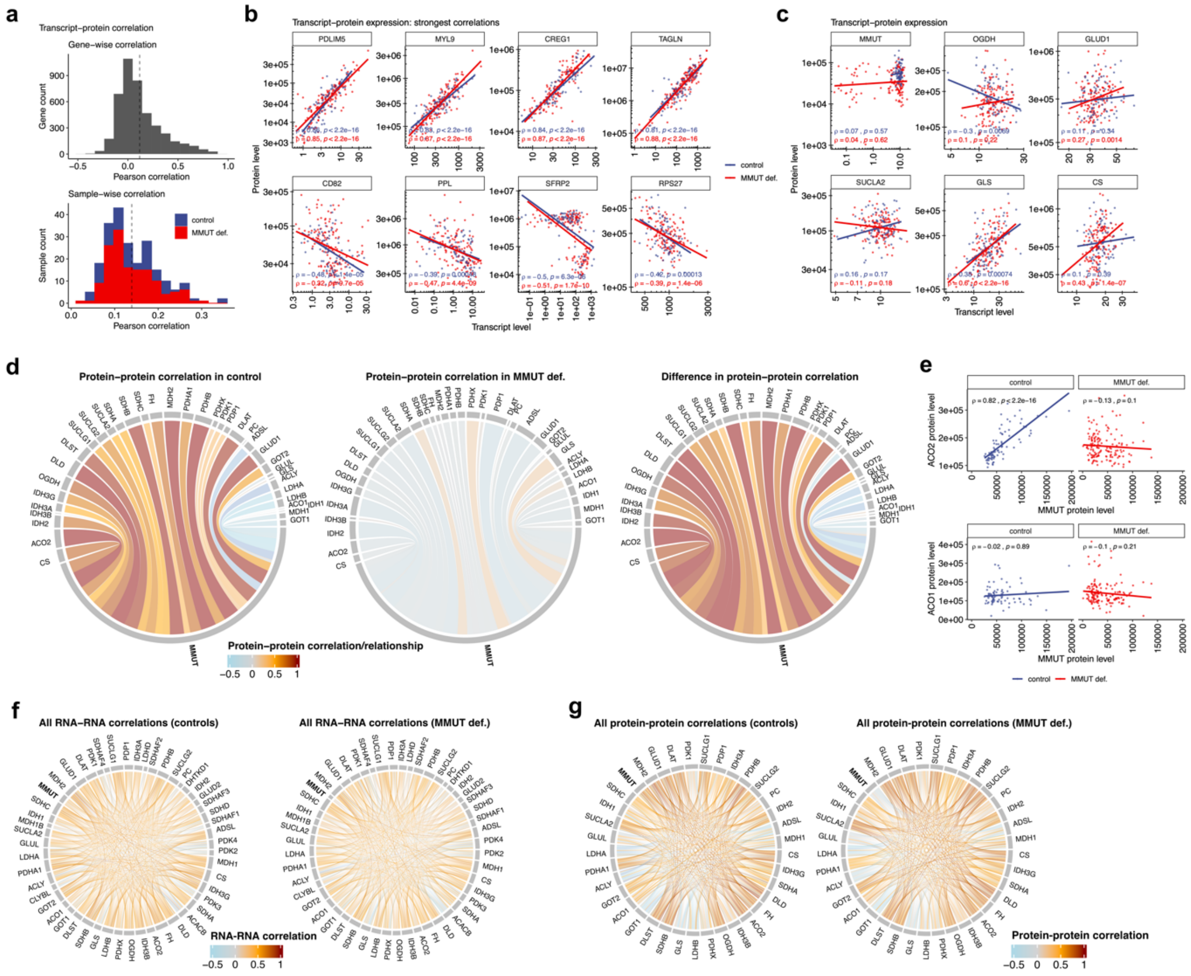

**a**, Histograms of Pearson correlations across 4318 transcript-protein pairs (top), and 221 samples (bottom). **b**, Scatter plot of the strongest positive and negative transcript-protein correlations (ranked by average of Spearman correlation coefficient in the MMUT-deficient and control group). **c**, Transcript-protein correlation plots of selected TCA cycle related genes. **d**, Spearman correlation of the MMUT protein versus a selection of TCA cycle and related proteins and their isoforms illustrated in a chord plot for control (left), MMUT-deficient samples (middle), and the difference of the two former plots (right); thickness of the links indicates nominal value of the correlation coefficient. **e**, Scatter plot of MMUT protein versus both isoforms of aconitase (ACO1 and ACO2) with linear regression by Spearman correlation. **f**, Chord plot illustrating all correlative relationships of TCA cycle and related transcripts or proteins (**g**); thickness of the links indicates nominal value of the Spearman correlation coefficient.

### Extended data Fig. 9 Metabolomics investigation of a subset of patient cell lines and glutamine flux studies in 293T CRISPR-KO cells.

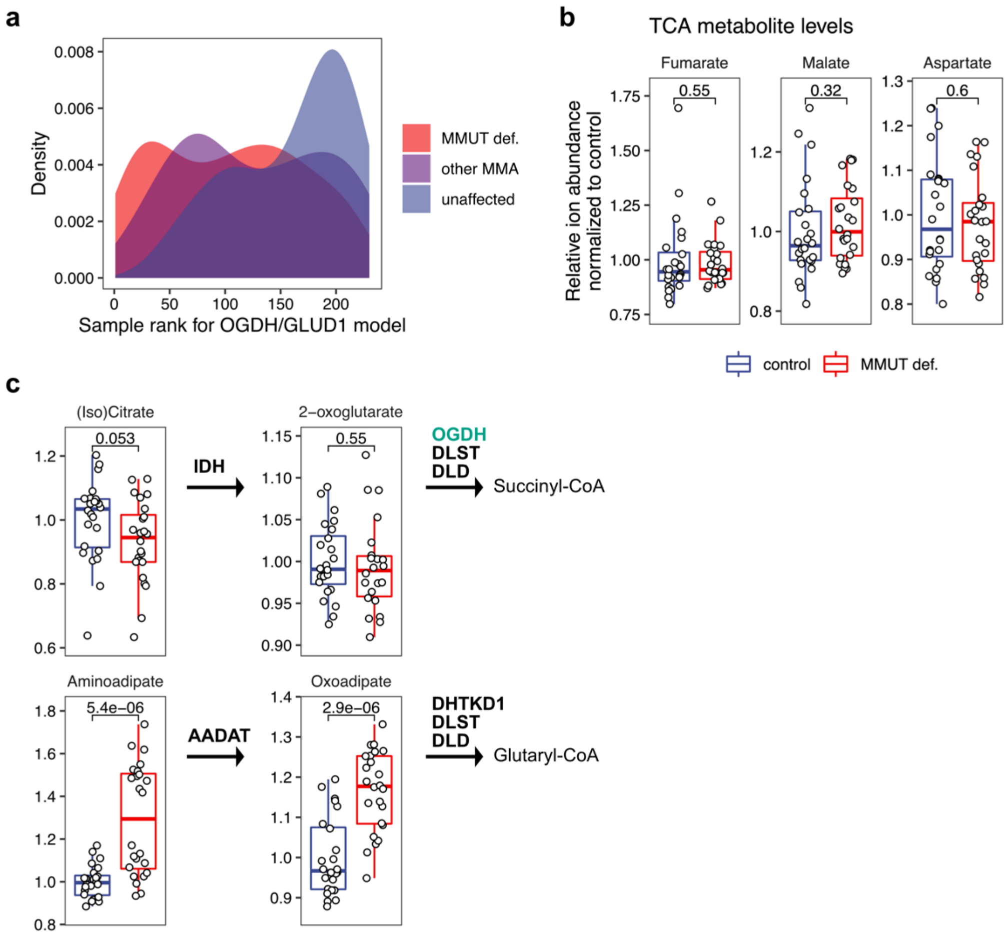

**a**, Density plot illustrating a model for OGDH and GLUD1 with all fibroblast samples ranked according to three sample groups. **b**, Non-significant changes of TCA metabolites as measured by untargeted polar metabolomics measured in a subset of patient-derived fibroblasts. **c**, Levels of metabolites involved in the two enzymatic steps catalyzed by two oxoacid dehydrogenase complexes (OGDC and OADC) and their proximal reactions; OGDH protein in green indicates its downregulation as detected in the proteotyping dataset.

#### Extended data Fig. 10 Validation of CRISPR knock-out 293T cell lines.

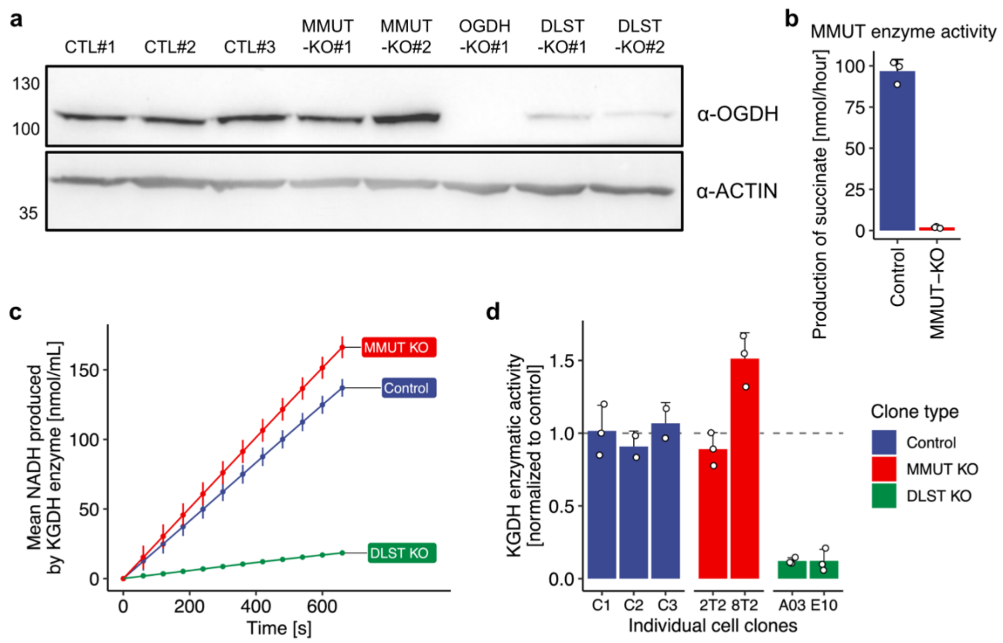

**a**, Western blots probing for OGDH in CRISPR knock-out cell lines. Cell line numbers indicate biological replicates; *OGDH*-KO cell line was not used in this study. **b**, MMUT enzymatic activity assessed by succinate production. **c**, Alpha-ketoglutarate dehydrogenase (KGDH) enzyme activity assessed by spectrophotometric measurement of produced NADH over time. **d**, Enzyme activities in individual clones normalized to the first control (wildtype) cell line.

#### Extended data Fig. 11 Stable isotope labeling of glutamine shows preferential reductive cycling of glutamine derived carbons in MMUT deficiency.

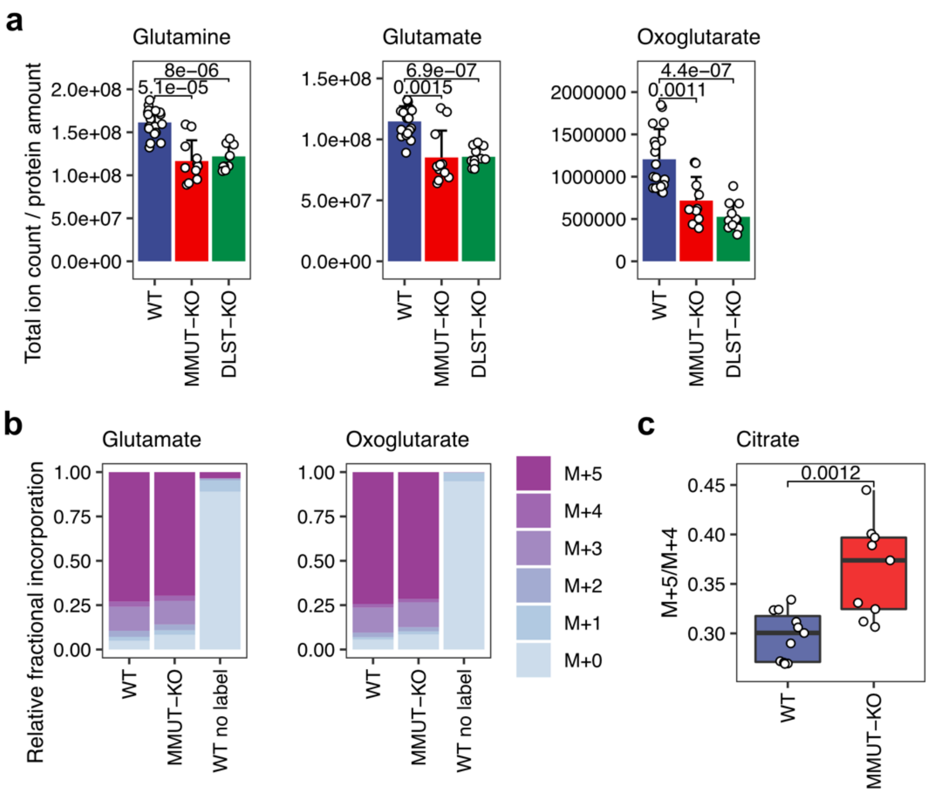

**a**, Pool sizes of metabolites in control and CRISPR/Cas9 KO 293T cells. **b**, Proportional bar plots illustrating the fractions of different isotopologues following labelling with [U-$^{13}$C]glutamine in 293T cells. **c**, M+5/M+4 ratio for citrate.

### Extended data Fig. 12 Immunoprecipitation of flag-tagged MMUT shows pulldown of DLST.

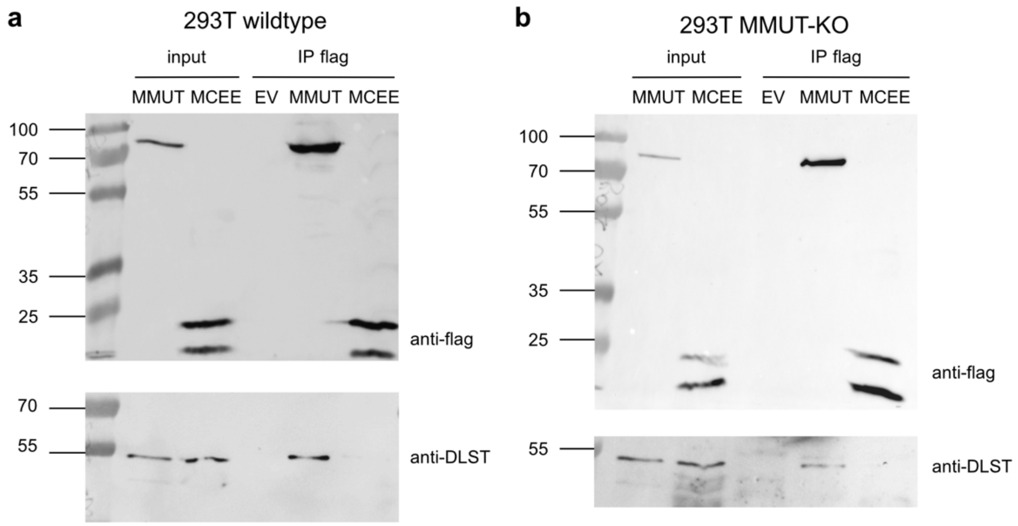

Western blot of immunoprecipitation of flag-tagged MMUT probing for DLST in 293T **a**, wildtype and **b**, *MMUT*-KO cell lines.

### Extended data Table 1 Overview of pathogenic variants.

Variants detected on WGS (novel variants in bold); nt, nucleotide; aa, amino acid.

| Cell line number | Forny et al. number (PMID: 27167370) | Gene | Variant 1 (nt) | Variant 1 (aa) | Variant 2 (nt) | Variant 2 (aa) | Comment |
|---|---|---|---|---|---|---|---|
| MMA001 | 2 | MMUT | c.654A>C | p.Gln218His | c.1106G>A | p.Arg369His | |
| MMA002 | 3 | MMUT | c.1106G>A | p.Arg369His | c.1106G>A | p.Arg369His | |
| MMA003 | 6 | MMUT | c.409C>T | **p.Ala137Val** | c.655A>T | p.Asn219Tyr | |
| MMA004 | 7 | MMUT | c.607G>A | p.Gly203Arg | c.1106G>A | p.Arg369His | |
| MMA005 | 8 | MMUT | c.982C>T | p.Leu328Phe | c.982C>T | p.Leu328Phe | |
| MMA006 | 9 | MMUT | c.1106G>A | p.Arg369His | c.691T>A | p.Tyr231Asn | |
| MMA007 | 10 | MMUT | c.1106G>A | p.Arg369His | c.1106G>A | p.Arg369His | |
| MMA008 | 11 | MMUT | c.1031C>T | **p.Ser344Phe** | c.312delC | p.Trp105Glyfs*75 | |
| MMA009 | 12 | MMUT | c.607G>A | p.Gly203Arg | c.607G>A | p.Gly203Arg | |
| MMA010 | 13 | MMUT | c.299A>G | p.Tyr100Cys | c.299A>G | p.Tyr100Cys | |
| MMA011 | 14 | MMUT | c.1361G>A | **p.Gly454Glu** | c.427C>T | **p.His143Tyr** | |
| MMA012 | 15.1 | MMUT | c.1106G>A | p.Arg369His | c.1097A>G | **p.Asn366Ser** | |
| MMA013 | 16 | MMUT | c.2081G>T | **p.Arg694Leu** | c.1207C>T | p.Arg403* | |
| MMA014 | 20 | MMUT | c.862T>C | **p.Ser288Pro** | c.862T>C | **p.Ser288Pro** | |
| MMA015 | 21 | MMUT | c.2080C>T | p.Arg694Trp | c.2080C>T | p.Arg694Trp | |
| MMA016 | 22 | MMUT | c.2099T>A | p.Met700Lys | c.623_624delTA | **p.Val208Alafs*2** | |
| MMA017 | 24 | MMUT | c.655A>T | p.Asn219Tyr | c.655A>T | p.Asn219Tyr | |
| MMA018 | 25 | MMUT | c.655A>T | p.Asn219Tyr | c.1420C>T | p.Arg474* | |
| MMA019 | 26 | MMUT | c.443C>T | **p.Ser148Leu** | c.1677-1G>C | Splice site | |
| MMA020 | 32 | MMUT | c.655A>T | p.Asn219Tyr | c.1560+1G>T | Splice site | |
| MMA021 | 33 | MMUT | c.655A>T | p.Asn219Tyr | c.1889G>A | p.Gly630Glu | |
| MMA022 | 34 | MMUT | c.1106G>A | p.Arg369His | c.2009delG | **p.Gly670Alafs*2** | |
| MMA023 | 36 | MMUT | c.572C>A | p.Ala191Glu | c.572C>A | p.Ala191Glu | |
| MMA024 | 38 | MMUT | c.2159-2160delAT | **p.Asn720Serfs*17** | c.2159-2160delAT | p.Asn720Serfs*736 | |
| MMA025 | 38.1 | MMUT | c.655A>T | p.Asn219Tyr | c.655A>T | p.Asn219Tyr | |
| MMA026 | 41 | MMUT | c.566A>T | p.Asn189Ile | c.1658delT | p.Val553Glyfs*17 | |
| MMA027 | 44 | MMUT | c.88C>T | p.Gln30* | c.88C>T | p.Gln30* | |
| MMA028 | 45 | MMUT | c.654A>C | p.Gln218His | c.654A>C | p.Gln218His | |
| MMA029 | 46 | MMUT | c.1962_1963delTC | p.Arg655* | c.1962_1963delTC | p.Arg655* | |
| MMA030 | 47 | MMUT | c.689C>G | p.Thr230Arg | c.689C>G | p.Thr230Arg | |
| MMA031 | 49 | MMUT | c.1084-10A>G | p.Gln361_Asp362insIlePhe* | c.1084-10A>G | p.Gln361_Asp362insIlePhe* | |
| MMA032 | 50 | MMUT | c.786T>G | **p.Ser262Arg** | c.1889G>A | p.Gly630Glu | |
| MMA033 | 51 | MMUT | c.-39-1G>A | splice site | c.-39-1G>A | Splice site | |
| MMA034 | 52 | MMUT | c.914T>C | p.Leu305Ser | c.914T>C | p.Leu305Ser | |
| MMA035 | 54 | MMUT | c.378C>A | p.Asn126Lys | c.974G>A | p.Gly325Asp | |
| MMA036 | 55 | MMUT | c.2179C>T | p.Arg727* | c.2179C>T | p.Arg727* | |
| MMA037 | 57 | MMUT | c.851G>A | p.Gly284Glu | c.982C>T | p.Leu328Phe | |
| MMA038 | 58 | MMUT | c.572C>A | p.Ala191Glu | c.1541delA | **p.Gln514Argfs*24** | |
| MMA039 | 59 | MMUT | c.1874A>T | **p.Asp625Val** | c.1874A>T | **p.Asp625Val** | |
| MMA040 | 60 | MMUT | c.421delG | p.Ala141Argfs*39 | c.421delG | p.Ala141Argfs*39 | |
| MMA041 | 61 | MMUT | c.1843C>A | p.Pro615Thr | c.2179C>T | p.Arg727* | |
| MMA042 | 62 | MMUT | c.692dupA | **p.Tyr231*** | c.692dupA | **p.Tyr231*** | |
| MMA043 | 63 | MMUT | c.982C>T | p.Leu328Phe | c.982C>T | p.Leu328Phe | |
| MMA044 | 67 | MMUT | c.753+2T>A | Splice site | c.2206C>T | p.Leu736Phe | |
| MMA045 | 74 | MMUT | c.1276G>A | p.Gly426Arg | c.1655C>T | p.Ala552Val | |
| MMA046 | 101 | MMUT | c.654A>C | p.Gln218His | c.1106G>A | p.Arg369His | |
| MMA047 | 121 | MMUT | c.1160C>T | **p.Thr387Ile** | c.1160C>T | **p.Thr387Ile** | |
| MMA048 | 125 | MMUT | c.655A>T | p.Asn219Tyr | c.828G>C | p.Glu276Asp | |
| MMA049 | 137 | MMUT | c.977G>A | p.Arg326Lys | c.2194_2197delinsTGGAA | p.Ala762Trpfs*6 | |
| MMA050 | 138 | MMUT | c.683G>A | **p.Arg228Gln** | c.2200C>T | p.Gln734* | |
| MMA051 | | MMUT | c.1022dupA | p.Asn341fs* | c.2150G>T | p.Gly717Val | |
| MMA052 | | MMUT | c.2080C>T | p.Arg694Trp | c.2080C>T | p.Arg694Trp | |

| | | | | | | |
|---|---|---|---|---|---|---|
| MMA053 | | MMUT | c.277C>T | p.Arg93Cys | c.1207C>T | p.Arg403* |
| MMA054 | | MMUT | c.91C>T | p.Arg31* | c.323G>A | p.Arg108His |
| MMA055 | | MMUT | c.662T>A | **p.Ile221Lys** | c.1790_1791insCT | **p.Thr598*** |
| MMA056 | | MMUT | c.1885A>G | p.Arg629Gly | c.1885A>G | p.Arg629Gly |
| MMA057 | | MMUT | **c.1560+3A>G** | Splice site | **c.1560+3A>G** | Splice site |
| MMA058 | | MMUT | c.850G>A | p.Gly284Arg | c.1073T>C | **p.Leu358Pro** |
| MMA059 | | MMUT | c.544dupA | **p.Met182Asnfs*29** | c.544dupA | **p.Met182Asnfs*29** |
| MMA060 | | MMUT | **c.1560+3A>G** | Splice site | **c.1560+3A>G** | Splice site |
| MMA061 | | MMUT | c.2179C>T | p.Arg727* | c.2179C>T | p.Arg727* |
| MMA062 | | MMUT | c.655A>T | p.Asn219Tyr | c.1055dupA | **p.Thr353Aspfs*** |
| MMA063 | | MMUT | c.1846C>T | p.Arg616Cys | c.1588_1595delGCTGAACG | **p.Ala530Leufs*11** |
| MMA064 | | MMUT | c.2080C>T | p.Arg694Trp | c.360dupT | p.Lys121* |
| MMA065 | | MMUT | c.-39-1G>A | splice site | c.-39-1G>A | Splice site |
| MMA066 | | MMUT | c.1531C>T | p.Arg511* | c.1531C>T | p.Arg511* |
| MMA067 | | MMUT | c.1843C>A | p.Pro615Thr | c.1843C>A | p.Pro615Thr |
| MMA068 | | MMUT | c.329A>G | p.Tyr110Cys | c.329A>G | p.Tyr110Cys |
| MMA069 | | MMUT | c.160A>T | p.Lys54* | c.160A>T | p.Lys54* |
| MMA070 | | MMUT | c.521T>C | p.Phe174Ser | c.521T>C | p.Phe174Ser |
| MMA071 | | MMUT | c.1181T>A | **p.Leu394*** | c.91C>T | p.Arg31* |
| MMA072 | | MMUT | c.1207C>T | p.Arg403* | c.572C>A | p.Ala191Glu |
| MMA073 | | MMUT | c.1808G>A | p.Arg603Lys | c.1808G>A | p.Arg603Lys |
| MMA074 | | MMUT | c.2115dupA | **p.Pro706Thrfs*6** | c.2115dupA | **p.Pro706Thrfs*6** |
| MMA075 | | MMUT | c.-39-1G>A | Splice site | c.91C>T | p.Arg31* |
| MMA076 | | MMUT | c.91C>T | p.Arg31* | c.91C>T | p.Arg31* |
| MMA077 | | MMUT | c.1106G>A | p.Arg369His | het deletion of exon 4 | |
| MMA078 | | MMUT | c.597T>C | p.Phe174Ser | c.597T>C | p.Phe174Ser |
| MMA079 | | MMUT | c.1889G>A | p.Gly630Glu | c.1889G>A | p.Gly630Glu |
| MMA080 | | MMUT | c.572C>A | p.Ala191Glu | c.572C>A | p.Ala191Glu |
| MMA081 | | MMUT | c.2179C>T | p.Arg727* | c.1106G>A | p.Arg369His |
| MMA082 | | MMUT | c.1531C>T | p.Arg511* | c.1531C>T | p.Arg511* |
| MMA083 | | MMUT | c.330T>G | p.Tyr110* | c.164delA | **p.Asn55Thrfs*5** |
| MMA084 | | MMUT | c.1962_1963delTC | p.Arg655* | c.1962_1963delTC | p.Arg655* |
| MMA085 | | MMUT | c.1690G>T | **p.Glu564*** | c.1690G>T | **p.Glu564*** |
| MMA086 | | MMUT | c.655A>T | p.Asn219Tyr | c.655A>T | p.Asn219Tyr |
| MMA087 | | MMUT | c.1844C>T | p.Pro615Leu | c.1844C>T | p.Pro615Leu |
| MMA088 | | | | | | |
| MMA089 | | MMUT | c.572C>A | p.Ala191Glu | c.2194_2197delinsTGGAA | p.Ala762Trpfs*6 |
| MMA090 | | MMUT | c.1843C>A | p.Pro615Thr | c.1843C>A | p.Pro615Thr |
| MMA091 | | MMUT | c.1240G>T | p.Glu414* | c.1240G>T | p.Glu414* |
| MMA092 | | MMUT | c.1207C>T | p.Arg403* | c.1207C>T | p.Arg403* |
| MMA093 | | MMUT | c.1311_1312insA | **p.Val438Serfs*3** | c.1311_1312insA | **p.Val438Serfs*3** |
| MMA094 | | MMUT | c.655A>T | p.Asn219Tyr | c.1782_1786delTAAAG | **p.Ser594Argfs*11** |
| MMA095 | | MMUT | c.655A>T | p.Asn219Tyr | c.322C>T | p.Arg108Cys |
| MMA096 | | MMUT | c.394C>T | p.Gln132* | c.394C>T | p.Gln132* |
| MMA097 | | MMUT | c.1843C>A | p.Pro615Thr | c.1843C>A | p.Pro615Thr |
| MMA098 | | MMUT | c.1880A>G | p.His627Arg | c.654A>C | p.Gln218His |
| MMA099 | | MMUT | c.647C>T | **p.Thr216Ile** | c.C647T | **p.Thr216Ile** |
| MMA100 | | MMUT | c.420C>T | p.Arg474* | c.753+2T>A | Splice site |
| MMA101 | | MMUT | c.1280G>A | p.Gly427Asp | c.323G>A | p.Arg108His |
| MMA102 | | MMUT | c.1280G>A | p.Gly427Asp | c.729_730insTT | p.Asp244Leufs* |
| MMA103 | | MMUT | c.654A>C | p.Gln218His | c.654A>C | p.Gln218His |
| MMA104 | | MMUT | c.454C>T | p.Arg152* | c.454C>T | p.Arg152* |
| MMA105 | | MMUT | c.884G>T | **p.Gly295Val** | c.884G>T | **p.Gly295Val** |
| MMA106 | | MMUT | **c.1560+3A>G** | Splice site | **c.1560+3A>G** | Splice site |
| MMA107 | | MMUT | c.1808G>A | p.Arg603Lys | c.1808G>A | p.Arg603Lys |
| MMA108 | | MMUT | c.454C>T | p.Arg152* | c.454C>T | p.Arg152* |

| | | | | | | | |
|---|---|---|---|---|---|---|---|
| MMA109 | | MMUT | c.1670G>C | **p.Arg557Pro** | c.1207C>T | p.Arg403* | |
| MMA110 | | MMUT | c.982C>T | p.Leu328Phe | c.360dupT | p.Lys121* | |
| MMA111 | | MMUT | c.146dupA | **p.Gln50Alafs*** | c.146dupA | **p.Gln50Alafs*** | |
| MMA112 | | MMUT | c.1399C>T | p.Arg467* | c.1399C>T | p.Arg467* | |
| MMA113 | | MMUT | c.1880A>G | p.His627Arg | c.655A>T | p.Asn219Tyr | |
| MMA114 | | MMUT | c.394C>T | p.Gln132* | c.323G>T | **p.Arg108Leu** | |
| MMA115 | | MMUT | c.160A>T | p.Lys54* | c.160A>T | p.Lys54* | |
| MMA116 | | MMUT | c.2080C>T | p.Arg694Trp | **c.754-5T>G** | Splice site | |
| MMA117 | | MMUT | c.1843C>A | p.Pro615Thr | c.1843C>A | p.Pro615Thr | |
| MMA118 | | MMUT | c.129G>A | p.Trp43* | c.129G>A | p.Trp43* | |
| MMA119 | | MMUT | c.88C>T | p.Gln30* | c.88C>T | p.Gln30* | |
| MMA120 | | MMUT | c.1399C>T | p.Arg467* | c.323G>A | p.Arg108His | |
| MMA121 | | MMUT | c.278G>A | p.Arg93His | c.278G>A | p.Arg93His | |
| MMA122 | | MMUT | c.129G>A | p.Trp43* | het deletion of exon 4 | | |
| MMA123 | | MMUT | c.655A>T | p.Asn219Tyr | c.655A>T | p.Asn219Tyr | |
| MMA124 | | MMUT | c.1846C>T | p.Arg616Cys | c.1846C>T | p.Arg616Cys | |
| MMA125 | | MMUT | c.1106G>A | p.Arg369His | c.1106G>A | p.Arg369His | |
| MMA126 | | | | | | | no coverage of the MMUT gene (homozygous deletion of 170 kB), clear cut to the surrounding regions which are well covered |
| MMA127 | | MMUT | c.1918G>T | **p.Asp640Tyr** | c.1912T>A | **p.Phe638Ile** | |
| MMA128 | | MMUT | c.C1843A | p.Pro615Thr | c.C1420T | p.Arg474* | |
| MMA129 | | MMUT | c.2T>C | p.Met1Thr | c.2T>C | p.Met1Thr | |
| MMA130 | | MMUT | c.682C>T | p.Arg228* | c.88C>T | p.Gln30* | |
| MMA131 | | MMUT | c.1677-1G>C | Splice site | c.1677-1G>C | Splice site | |
| MMA132 | | MMUT | c.422C>A | p.Ala141Glu | c.323G>A | p.Arg108His | |
| MMA133 | | MMUT | c.421delG | p.Ala141Argfs*39 | c.421delG | p.Ala141Argfs*39 | |
| MMA134 | | MMUT | c.360dupT | p.Lys121* | c.360dupT | p.Lys121* | |
| MMA135 | | MMUT | c.323G>A | p.Arg108His | c.1758delA | **p.Tyr587Ilefs*11** | |
| MMA136 | | MMUT | c.1843C>A | p.Pro615Thr | c.1843C>A | p.Pro615Thr | |
| MMA137 | | MMUT | c.643G>A | p.Gly215Ser | c.454C>T | p.Arg152* | |
| MMA138 | | MMUT | c.1106G>A | p.Arg369His | c.1560+1G>T | Splice site | |
| MMA139 | | | | | | | |
| MMA140 | | MMUT | c.278G>A | p.Arg93His | c.278G>A | p.Arg93His | |
| MMA141 | | MMUT | c.1207C>T | p.Arg403* | c.1207C>T | p.Arg403* | |
| MMA142 | | MMUT | c.1843C>A | p.Pro615Thr | c.1843C>A | p.Pro615Thr | |
| MMA143 | | MMUT | c.1038_1040delTCT | p.Leu347del | c.1038_1040delTCT | p.Leu347del | |
| MMA144 | | MMUT | c.397G>A | p.Gly133Arg | c.397G>A | p.Gly133Arg | |
| MMA145 | | MMUT | c.572C>A | p.Ala191Glu | c.572C>A | p.Ala191Glu | |
| MMA146 | | MMUT | c.1105C>T | p.Arg369Cys | c.91C>T | p.Arg31* | |
| MMA147 | | MMUT | c.2080C>T | p.Arg694Trp | c.1677-1G>C | Splice site | |
| MMA148 | | MMUT | **c.1444+2T>G** | Splice site | c.839dupC | **p.Leu281Phefs*9** | |
| MMA149 | | MMUT | c.1240G>T | p.Glu414* | c.1240G>T | p.Glu414* | |
| MMA150 | | MMUT | c.1962_1963delTC | p.Arg655* | c.1962_1963delTC | p.Arg655* | |
| MMA151 | | ACSF3 | c.1066G>A | p.Gly356Ser | c.1066G>A | p.Gly356Ser | |
| MMA152 | | MMAB | expression outlier | | | | cblB by complementation |
| MMA153 | | | | | | | |
| MMA154 | | ACSF3 | c.401T>C | p.Leu134Pro | c.401T>C | p.Leu134Pro | |
| MMA155 | | ACSF3 | c.1412G>A | p.Arg471Gln | c.1412G>A | p.Arg471Gln | |
| MMA156 | | ACSF3 | c.1A>G | p.Met1Val | c.1A>G | p.Met1Val | |
| MMA157 | | | | | | | |
| MMA158 | | ACSF3 | c.1672C>T | p.Arg558Trp | | | |
| MMA159 | | | | | | | |
| MMA160 | | ACSF3 | c.1672C>T | p.Arg558Trp | c.1075G>A | p.Glu359Lys | |
| MMA161 | | | | | | | |
| MMA162 | | | | | | | |
| MMA163 | | | | | | | |
| MMA164 | | SUCLA2 | c.534+1G>A | Splice site | c.534+1G>A | Splice site | |
| MMA165 | | ACSF3 | c.1470G>C | p.Glu490Asp | c.1470G>C | p.Glu490Asp | |

| | | | | | | | |
|---|---|---|---|---|---|---|---|
| MMA166 | | | | | | | |
| MMA167 | | | | | | | |
| MMA168 | | | | | | | |
| MMA169 | | | | | | | |
| MMA170 | | ACSF3 | c.1543C>T | p.Arg515Trp | c.1672C>T | p.Arg558Trp | |
| MMA171 | | | | | | | |
| MMA172 | | | | | | | |
| MMA173 | | ACSF3 | c.1614-2A>G | Splice site | c.1613+3A>C | splice site | |
| MMA174 | | | | | | | |
| MMA175 | | ACSF3 | c.1412G>A | p.Arg471Gln | c.1412G>A | p.Arg471Gln | |
| MMA176 | | TCN2 | c.172delC | p.Leu58Tyrfs *28 | c.172delC | p.Leu58Tyrfs *28 | |
| MMA177 | | | | | | | |
| MMA178 | | | | | | | |
| MMA179 | | ACSF3 | c.1470G>C | p.Glu490Asp | c.1470G>C | p.Glu490Asp | |
| MMA180 | | | | | | | |
| MMA181 | | | | | | | |
| MMA182 | | ACSF3 | c.1672C>T | p.Arg558Trp | c.1672C>T | p.Arg558Trp | |
| MMA183 | | SUCLA2 | c.664-1G>A | Splice site | c.664-1G>A | Splice site | |
| MMA184 | | | | | | | |
| MMA185 | | | | | | | |
| MMA186 | | ACSF3 | expression outlier | | | | |
| MMA187 | | ACSF3 | expression outlier | | | | |
| MMA188 | | | | | | | |
| MMA189 | | ACSF3 | c.1672C>T | p.Arg558Trp | c.1075G>A | p.Glu359Lys | |
| MMA190 | | | | | | | |
| MMA191 | | | | | | | |
| MMA192 | | SUCLA2 | c.1106dupA | p.Val370Glyf s*16 | | | |
| MMA193 | | ACSF3 | c.1470G>C | p.Glu490Asp | c.1470G>C | p.Glu490Asp | |
| MMA194 | | | | | | | |
| MMA195 | | TCN2 | c.328dupC | p.Leu110Prof s*8 | c.328dupC | p.Leu110Prof s*8 | |
| MMA196 | | ACSF3 | c.1672C>T | p.Arg558Trp | c.1672C>T | p.Arg558Trp | |
| MMA197 | | ACSF3 | c.1672C>T | p.Arg558Trp | c.1672C>T | p.Arg558Trp | |
| MMA198 | | TCN2 | c.497_498del TC | p.Leu166Prof s*7 | c.1137_1138i nsA | p.Tyr380Ilefs *32 | |
| MMA199 | | | | | | | |
| MMA200 | | | | | | | |
| MMA201 | | | | | | | |
| MMA202 | | ACSF3 | c.1672C>T | p.Arg558Trp | c.1672C>T | p.Arg558Trp | |
| MMA203 | | ACSF3 | c.1672C>T | p.Arg558Trp | c.1672C>T | p.Arg558Trp | |
| MMA204 | | | | | | | |
| MMA205 | | | | | | | |
| MMA206 | | MMAA | expression outlier | | | | cblA by complementation |
| MMA207 | | | | | | | |
| MMA208 | | | | | | | |
| MMA209 | | MMAA | expression outlier | | | | MMAA cDNA transcript not amplifiable |
| MMA210 | | | | | | | cblB by complementation |
| MMA211 | | | | | | | |
| MMA212 | | | | | | | |
| MMA213 | | | | | | | |
| MMA214 | | | | | | | |
| MMA215 | | | | | | | |
| MMA216 | | | | | | | |
| MMA217 | | | | | | | |
| MMA218 | | | | | | | |
| MMA219 | | | | | | | |
| MMA220 | | | | | | | |
| MMA221 | | | | | | | |
| MMA222 | | | | | | | |
| MMA223 | | | | | | | |
| MMA224 | | | | | | | |
| MMA225 | | | | | | | |
| MMA226 | | | | | | | |
| MMA227 | | | | | | | |

| |
|---|
| MMA228 |
| MMA229 |
| MMA230 |

### Extended data Table 2 List of significantly enriched proteins in pull-down by affinity capture mass spectrometry using MMUT, MMAB, or MCEE as baits.

| Accession Number | Molecular Weight | ANOVA Test (p-value) | Quantitative Profile |
|---|---|---|---|
| **MMUT-pulldown** | | | |
| AATM_HUMAN | 48 kDa | 0.0001 | EV_flag low, MUT_flag high, VLCAD_flag low |
| CH10_BOVIN (+1) | 11 kDa | 0.00022 | EV_flag low, MUT_flag high, VLCAD_flag low |
| ODPB_HUMAN | 39 kDa | 0.0012 | EV_flag low, MUT_flag high, VLCAD_flag low |
| MUTA_HUMAN [3] | 83 kDa | 0.0014 | EV_flag low, MUT_flag high, VLCAD_flag low |
| GRPE1_HUMAN (+1) | 24 kDa | 0.0039 | EV_flag low, MUT_flag high, VLCAD_flag low |
| P5CR1_HUMAN | 33 kDa | 0.0039 | EV_flag low, MUT_flag high, VLCAD_flag low |
| ATP5H_HUMAN | 18 kDa | 0.0039 | EV_flag low, MUT_flag high, VLCAD_flag low |
| EFTU_HUMAN | 50 kDa | 0.0042 | EV_flag low, MUT_flag high, VLCAD_flag low |
| P5CS_HUMAN | 87 kDa | 0.013 | EV_flag low, MUT_flag high, VLCAD_flag low |
| MDHM_HUMAN | 36 kDa | 0.015 | EV_flag low, MUT_flag high, VLCAD_flag low |
| SSBP_HUMAN | 17 kDa | 0.019 | EV_flag low, MUT_flag high, VLCAD_flag low |
| GLYM_HUMAN [2] | 56 kDa | 0.021 | EV_flag low, MUT_flag high, VLCAD_flag low |
| ATPO_HUMAN | 23 kDa | 0.022 | EV_flag low, MUT_flag high, VLCAD_flag low |
| RT23_HUMAN | 22 kDa | 0.027 | EV_flag low, MUT_flag high, VLCAD_flag low |
| RT27_HUMAN | 48 kDa | 0.027 | EV_flag low, MUT_flag high, VLCAD_flag low |
| STML2_HUMAN | 39 kDa | 0.031 | EV_flag low, MUT_flag high, VLCAD_flag low |
| ATPG_HUMAN | 33 kDa | 0.031 | EV_flag low, MUT_flag high, VLCAD_flag low |
| ODO2_HUMAN | 49 kDa | 0.034 | EV_flag low, MUT_flag high, VLCAD_flag low |
| IDH3A_HUMAN | 40 kDa | 0.034 | EV_flag low, MUT_flag high, VLCAD_flag low |
| ETFB_HUMAN | 28 kDa | 0.046 | EV_flag low, MUT_flag high, VLCAD_flag low |
| C1QBP_HUMAN | 31 kDa | 0.046 | EV_flag low, MUT_flag high, VLCAD_flag low |
| DHE3_HUMAN [2] | 61 kDa | 0.048 | EV_flag low, MUT_flag high, VLCAD_flag low |
| **MMAB-pulldown** | | | |
| MMAB_HUMAN | 27 kDa | 0.0001 | EV_flag low, MMAB_flag high, VLCAD_flag low |
| CH10_BOVIN (+1) | 11 kDa | 0.0001 | EV_flag low, MMAB_flag high, VLCAD_flag low |
| DHE3_HUMAN [2] | 61 kDa | 0.00029 | EV_flag low, MMAB_flag high, VLCAD_flag low |
| ATPO_HUMAN | 23 kDa | 0.00069 | EV_flag low, MMAB_flag high, VLCAD_flag low |
| P5CR1_HUMAN | 33 kDa | 0.0012 | EV_flag low, MMAB_flag high, VLCAD_flag low |
| ETFB_HUMAN | 28 kDa | 0.0018 | EV_flag low, MMAB_flag high, VLCAD_flag low |
| SYDM_HUMAN | 74 kDa | 0.0024 | EV_flag low, MMAB_flag high, VLCAD_flag low |
| ATPB_HUMAN [45] | 57 kDa | 0.0037 | EV_flag low, MMAB_flag high, VLCAD_flag low |
| ODPB_HUMAN | 39 kDa | 0.0039 | EV_flag low, MMAB_flag high, VLCAD_flag low |
| C1TM_HUMAN | 106 kDa | 0.0039 | EV_flag low, MMAB_flag high, VLCAD_flag low |
| HCDH_HUMAN | 34 kDa | 0.0039 | EV_flag low, MMAB_flag high, VLCAD_flag low |
| ALDH2_HUMAN | 56 kDa | 0.0039 | EV_flag low, MMAB_flag high, VLCAD_flag low |
| EFTU_HUMAN | 50 kDa | 0.0044 | EV_flag low, MMAB_flag high, VLCAD_flag low |
| MDHM_HUMAN | 36 kDa | 0.0053 | EV_flag low, MMAB_flag high, VLCAD_flag low |
| AATM_HUMAN | 48 kDa | 0.0076 | EV_flag low, MMAB_flag high, VLCAD_flag low |
| ECHB_HUMAN | 51 kDa | 0.0076 | EV_flag low, MMAB_flag high, VLCAD_flag low |
| TRAP1_HUMAN [2] | 80 kDa | 0.0085 | EV_flag low, MMAB_flag high, VLCAD_flag low |
| TRAP1_HUMAN | 80 kDa | 0.0078 | EV_flag low, MMAB_flag high, VLCAD_flag low |
| THIL_HUMAN | 45 kDa | 0.013 | EV_flag low, MMAB_flag high, VLCAD_flag low |
| CISY_HUMAN | 52 kDa | 0.014 | EV_flag low, MMAB_flag high, VLCAD_flag low |
| SSBP_HUMAN | 17 kDa | 0.014 | EV_flag low, MMAB_flag high, VLCAD_flag low |
| P5CS_HUMAN | 87 kDa | 0.02 | EV_flag low, MMAB_flag high, VLCAD_flag low |
| RT23_HUMAN | 22 kDa | 0.022 | EV_flag low, MMAB_flag high, VLCAD_flag low |
| GLYM_HUMAN [2] | 56 kDa | 0.025 | EV_flag low, MMAB_flag high, VLCAD_flag low |
| QCR1_HUMAN | 53 kDa | 0.027 | EV_flag low, MMAB_flag high, VLCAD_flag low |
| ATPA_HUMAN [2] | 60 kDa | 0.031 | EV_flag low, MMAB_flag high, VLCAD_flag low |
| ATPG_HUMAN | 33 kDa | 0.031 | EV_flag low, MMAB_flag high, VLCAD_flag low |
| GRP75_HUMAN | 74 kDa | 0.033 | EV_flag low, MMAB_flag high, VLCAD_flag low |
| CH60_HUMAN [2] | 61 kDa | 0.037 | EV_flag low, MMAB_flag high, VLCAD_flag low |
| **MCEE-pulldown** | | | |
| THIL_HUMAN | 45 kDa | 0.00048 | EV_flag low, MCEE_flag high, VLCAD_flag low |
| MCEE_HUMAN | 19 kDa | 0.0012 | EV_flag low, MCEE_flag high, VLCAD_flag low |
| ODO2_HUMAN | 49 kDa | 0.0012 | EV_flag low, MCEE_flag high, VLCAD_flag low |
| CALX_HUMAN | 68 kDa | 0.0012 | EV_flag low, MCEE_flag high, VLCAD_flag low |
| ODPB_HUMAN | 39 kDa | 0.0039 | EV_flag low, MCEE_flag high, VLCAD_flag low |
| THIM_HUMAN | 42 kDa | 0.0039 | EV_flag low, MCEE_flag high, VLCAD_flag low |
| HCD2_HUMAN | 27 kDa | 0.0052 | EV_flag low, MCEE_flag high, VLCAD_flag low |

| | | | |
|---|---|---|---|
| GRPE1_HUMAN (+1) | 24 kDa | 0.0052 | EV_flag low, MCEE_flag high, VLCAD_flag low |
| ECHB_HUMAN | 51 kDa | 0.0052 | EV_flag low, MCEE_flag high, VLCAD_flag low |
| ACADM_HUMAN (+1) | 47 kDa | 0.0076 | EV_flag low, MCEE_flag high, VLCAD_flag low |
| PGAM5_HUMAN | 32 kDa | 0.008 | EV_flag low, MCEE_flag high, VLCAD_flag low |
| RPN1_HUMAN | 69 kDa | 0.008 | EV_flag low, MCEE_flag high, VLCAD_flag low |
| ALDH2_HUMAN | 56 kDa | 0.008 | EV_flag low, MCEE_flag high, VLCAD_flag low |
| GLYM_HUMAN [2] | 56 kDa | 0.0088 | EV_flag low, MCEE_flag high, VLCAD_flag low |
| ETFB_HUMAN | 28 kDa | 0.0091 | EV_flag low, MCEE_flag high, VLCAD_flag low |
| MDHM_HUMAN | 36 kDa | 0.011 | EV_flag low, MCEE_flag high, VLCAD_flag low |
| ATPG_HUMAN | 33 kDa | 0.011 | EV_flag low, MCEE_flag high, VLCAD_flag low |
| EFTU_HUMAN | 50 kDa | 0.013 | EV_flag low, MCEE_flag high, VLCAD_flag low |
| CH10_BOVIN (+1) | 11 kDa | 0.015 | EV_flag low, MCEE_flag high, VLCAD_flag low |
| AATM_HUMAN | 48 kDa | 0.016 | EV_flag low, MCEE_flag high, VLCAD_flag low |
| DHE3_HUMAN [2] | 61 kDa | 0.019 | EV_flag low, MCEE_flag high, VLCAD_flag low |
| CH60_HUMAN [2] | 61 kDa | 0.019 | EV_flag low, MCEE_flag high, VLCAD_flag low |
| ATD3A_HUMAN [2] | 71 kDa | 0.021 | EV_flag low, MCEE_flag high, VLCAD_flag low |
| SSBP_HUMAN | 17 kDa | 0.021 | EV_flag low, MCEE_flag high, VLCAD_flag low |
| ATPA_HUMAN [2] | 60 kDa | 0.022 | EV_flag low, MCEE_flag high, VLCAD_flag low |
| ATPO_HUMAN | 23 kDa | 0.025 | EV_flag low, MCEE_flag high, VLCAD_flag low |
| ATPB_HUMAN [45] | 57 kDa | 0.026 | EV_flag low, MCEE_flag high, VLCAD_flag low |
| RL34_HUMAN | 13 kDa | 0.027 | EV_flag low, MCEE_flag high, VLCAD_flag low |
| TRAP1_HUMAN [2] | 80 kDa | 0.028 | EV_flag low, MCEE_flag high, VLCAD_flag low |
| TRAP1_HUMAN | 80 kDa | 0.028 | EV_flag low, MCEE_flag high, VLCAD_flag low |
| RT23_HUMAN | 22 kDa | 0.031 | EV_flag low, MCEE_flag high, VLCAD_flag low |
| SDHA_HUMAN (+1) | 73 kDa | 0.031 | EV_flag low, MCEE_flag high, VLCAD_flag low |
| SSDH_HUMAN | 57 kDa | 0.031 | EV_flag low, MCEE_flag high, VLCAD_flag low |
| ODP2_HUMAN | 69 kDa | 0.034 | EV_flag low, MCEE_flag high, VLCAD_flag low |
| OAT_HUMAN | 49 kDa | 0.034 | EV_flag low, MCEE_flag high, VLCAD_flag low |
| ATP5H_HUMAN | 18 kDa | 0.034 | EV_flag low, MCEE_flag high, VLCAD_flag low |
| IPYR2_HUMAN | 38 kDa | 0.034 | EV_flag low, MCEE_flag high, VLCAD_flag low |
| CISY_HUMAN | 52 kDa | 0.05 | EV_flag low, MCEE_flag high, VLCAD_flag low |
